# Innate lymphoid cells are activated and their levels correlate with viral load in patients with Puumala hantavirus caused hemorrhagic fever with renal syndrome

**DOI:** 10.1101/2022.05.10.22274837

**Authors:** Marina García, Anna Carrasco García, Johanna Tauriainen, Kimia Maleki, Antti Vaheri, Satu Mäkelä, Jukka Mustonen, Anna Smed-Sörensen, Tomas Strandin, Jenny Mjösberg, Jonas Klingström

## Abstract

**Background:** Innate lymphoid cells (ILCs) are involved in immunity and homeostasis but, except for natural killer (NK) cells, their role in human viral infections is not well known. Puumala virus (PUUV) is a hantavirus that causes the acute zoonotic disease hemorrhagic fever with renal syndrome (HFRS). HFRS is characterized by strong systemic inflammation and NK cells are highly activated in HFRS, suggesting that also other ILCs might be responding to infection.

**Methods:** Here we phenotypically analyzed peripheral ILCs in acute and convalescent PUUV-infected HFRS patients. Additionally, plasma levels of soluble factors and viral load were analyzed.

**Findings:** Overall, the frequencies of NK cells and naïve ILCs were reduced while the frequency of ILC2, in particular the ILC2-lineage committed c-Kit^lo^ ILC2 subset, was increased during acute HFRS. Interestingly, we observed a negative correlation between viral load and frequencies of both NK and non-NK ILCs in acute HFRS. Phenotypically, ILCs displayed an activated profile with increased proliferation, and showed altered expression of several homing markers during acute HFRS. In line with the observation of activated ILCs, plasma levels of inflammatory proteins, including the ILC-associated cytokines interleukin (IL)-13, IL-23, IL-25, IL-33, and thymic stromal lymphopoietin (TSLP), were elevated during acute HFRS.

**Interpretation:** These findings indicate a general involvement of ILCs in response to human hantavirus infection. Further, this constitutes the first comprehensive study of ILCs in a hantavirus-caused disease, aiding in further understanding the role of these cells in disease pathogenesis and in human viral infections in general.

**Funding:** A full list of funding bodies that contributed to this study can be found in the Acknowledgements section.

**RESEARCH IN CONTEXT:** 

**Evidence before this study:** Innate lymphoid cells (ILCs) include a broad range of innate cell subsets involved, among other functions, in early response to infections. Natural killer (NK) cells have been broadly characterized in human viral infections, but much less is known in such context about other more recently discovered ILCs. Puumala virus is one of the causative agents of hemorrhagic fever with renal syndrome (HFRS), an acute zoonotic disease characterized by systemic inflammation. No current treatment or vaccines are available to date for hantavirus-caused diseases and pathogenesis is still not fully understood. A PubMed search up until April 2022 using a combination of the terms virus, Puumala virus, hantavirus, HFRS, ILCs, and NK cells shows that while studies have been performed characterizing NK cells in HFRS, no studies are available on the rest of ILCs in hantavirus-caused diseases, which in addition have been studied in only a handful of human viral infections.

**Added value of this study:** In this study we thoroughly characterized the ILC landscape in circulation and its milieu in acute and convalescent Puumala-infected HFRS patients. We found that the frequency of NK cells and other ILC subsets was altered in the acute HFRS patients as compared to convalescent HFRS patients and control individuals, with a decrease in NK cells and naïve ILCs, and an increase in ILC2. In particular the ILC2-lineage committed c-Kit^lo^ ILC2 subset was found increased. Total ILCs showed increased levels of activation and proliferation, and signs of altered migration patterns in acute HFRS patients. The finding of elevated levels of several soluble inflammatory proteins associated with ILCs in acute HFRS patients further suggests an implication of ILCs in HFRS. Moreover, an association between the frequency of NK cells and non-NK ILCs and plasma viral load suggest a possible connection between viremia and the activity of these cell types during the course of disease.

**Implications of all the available evidence:** Our research shows that all circulating ILCs, including non-NK ILCs, are activated, proliferating, and correlate with viral load in acute HFRS patients, indicating a general involvement in human hantavirus infection and disease. Being the first comprehensive study on ILCs in hantavirus infections, further research will give relevant insight into the roles of ILCs in disease pathogenesis and protection.

## INTRODUCTION

Innate lymphoid cells (ILCs) are a group of innate immune cells that play important roles in the modulation of immune and inflammatory responses.^1^ Naïve ILCs (nILC) constitute an immature subset ^2,3^ that can home from peripheral blood to tissues where they give rise to the mature ILC subsets ^4^. Mature ILCs are classified in five main subsets based on the transcription factors they express and the cytokines they produce: natural killer (NK) cells, ILC1, ILC2, ILC3, and lymphoid tissue inducer (LTi) cells.^5,6^ NK cells share the same features as ILC1, but in addition can, as opposed to the other ILCs, kill virus-infected cells.^6^ ILC1 and ILC3 are mainly found in mucosal tissues while NK cells and ILC2 are found both in tissues and in peripheral blood.^2,4^ Furthermore, two functionally distinct subsets of ILC2 can be found in peripheral blood: c-Kit^lo^ and c-Kit^hi^ ILC2, the first being more committed to the ILC2 lineage.^7^

NK cells have been extensively described in different viral infections.^8–10^ On the other hand, non-NK ILCs (hereinafter referred to as ILCs) were more recently discovered,^11,12^ and thus less is known regarding their role in viral infections. Due to their location in mucosal tissue, ILCs are on the first line of defense and hence potentially essential in the early phases of viral infections.^13^ Their enrichment in lungs suggests an important role for ILCs in respiratory viral infections.^13,14^ There are conflicting data regarding ILC2 and virus infections: they have been suggested to promote tissue repair and protect the lungs of influenza-infected mice,^15,16^ while other studies reported that ILC2 induce airway hyperreactivity in influenza-infected ^17–19^ and respiratory syncytial virus-infected ^20^ mice. Furthermore, ILCs have recently been investigated in the context of a few human viral diseases. In HIV-1 infected individuals, levels of ILCs were found to be reduced in ileum and colon, ^21^ as well as in circulation, and to negatively correlate with viral load.^22^ Total peripheral ILC levels were also found to be decreased in SARS-CoV-2-infected coronavirus disease-19 (COVID-19) patients, with ILC2 decreased in severe but not in moderate patients.^23–25^ Moreover, in infants with respiratory syncytial virus bronchiolitis, elevated levels of ILC2 in the airways were found to associate with disease severity.^26^ Overall, these studies show an effect of viral infections on the ILC landscape.

Hantaviruses (genus *Orthohantavirus*, family *Hantaviridae*, order *Bunyavirales*) are RNA viruses that can cause zoonotic diseases in humans ^27^. The reservoirs of disease-causing hantaviruses are rodents, and humans are normally infected via inhalation of hantavirus-infected rodent excreta.^28,29^ Hantaviruses can cause hemorrhagic fever with renal syndrome (HFRS) in Eurasia and hantavirus pulmonary syndrome (HPS) in the Americas, with up to 10% and around 35% case fatality rate, respectively.^28,30,31^ Puumala virus (PUUV) is endemic in Europe, where it is the most common causative agent of HFRS.^30,32^ Hantavirus-infected patients initially develop non-specific symptoms such as high fever and headache, and many eventually present kidney dysfunction (primarily in HFRS), lung dysfunction (primarily in HPS), and gastrointestinal symptoms, including abdominal pain, diarrhea, vomiting, and gastrointestinal bleeding.^31,33,34^ As of today, no specific treatment nor United States Food and Drug Administration / European Medicines Agency-approved vaccines are available for hantavirus-caused diseases.^35^ Hantaviruses trigger immunopathogenic responses that likely contribute to hyperinflammation ^36–43^ and vascular leakage in patients, but the exact mechanisms leading to these events remain to be understood.^32,44,45^ Peripheral NK cells show signs of strong activation and proliferation in the acute phase of HFRS.^46–49^ B and T cells are highly expanded in the circulation of HFRS and HPS patients, with concomitant elevated levels of CD8^+^ T cells in the respiratory airways, ^36,50–54^. Mucosa-associated invariant T (MAIT) cells were recently shown to be reduced but highly activated in circulation during PUUV-caused HFRS.^42^ Levels of neutrophils are highly increased and also strongly activated in hantavirus-infected patients.^55–57^ Mononuclear phagocytes are susceptible to infection with hantaviruses, and their activation and redistribution from circulation towards the airways and kidneys in HFRS patients have been reported.^58–61^ Combined, these reports show that hantaviruses trigger strong immune cell responses, indicating a possible involvement of ILCs as well.

Here we performed a detailed characterization of peripheral blood ILCs and NK cells, as well as of their cytokine and chemokine milieu, in PUUV-infected HFRS patients. We observed increased plasma levels of inflammatory proteins, including ILC-associated cytokines. We showed that NK cell frequencies are reduced during acute HFRS but recover during convalescence. While total ILC frequencies did not change, we report increased frequency of ILC2 and a concomitant decreased frequency of nILC during acute HFRS. In particular, the ILC2-lineage committed c-Kit^lo^ ILC2 subset was increased during acute HFRS. Furthermore, NK cells and ILCs displayed an activated phenotype and ongoing proliferation during acute HFRS. Interestingly, we observed a negative correlation between viral load and the frequencies of both NK cells and ILCs in acute HFRS, suggesting a potential direct or indirect influence of hantaviruses on the ILC landscape in HFRS patients.

## MATERIAL AND METHODS

### Patient samples

17 patients with serologically confirmed acute HFRS were included in the study. The patients were diagnosed with PUUV infection at the Tampere University Hospital, Finland during 2002–2007. Whole blood samples were collected, and peripheral blood mononuclear cells (PBMCs) were isolated as previously described and stored in liquid nitrogen at -150°C, while plasma was stored at -80°C, until further use. ^61^

The study was approved by the Ethics Committee of Tampere University Hospital (ethical permit nr. R04180) and all subjects gave written informed consent. The samples were collected in the acute phase of disease (5-8 days after onset of symptoms), during the early convalescence phase (20-27 days after onset of symptoms), and at a late convalescence phase at 180 or 360 days after onset of symptoms. Patients were stratified as having either mild or severe HFRS based on a scoring system adapted from the sequential organ failure assessment scoring system, where the maximum levels of creatinine (4 = > 440, 3 = 300-440, 2 = 171-299, 1 = 110-170, and 0 = < 110 µmol/l), minimum level of platelets (4 = < 20, 3 = 20-49, 2 = 50-99, 1 = 100-150, and 0 = > 150 x 10^3^/µl), and minimum mean arterial blood pressure (1 = < 70 and 0 = ≥ 70 mmHg) were ranked. A total score of ≥ 5 was considered severe and < 5 mild. ^61,62^

As controls, PBMCs were obtained from buffy coats from 10 healthy blood donors from the Blood Transfusion Clinic at the Karolinska University Hospital Huddinge, Stockholm, Sweden (ethical permit nr. 2020-02604) and stored in liquid nitrogen until further use. PBMCs were isolated from the buffy coats by density centrifugation using Lymphoprep (StemCell Technologies), according to manufacturer’s guidelines.

### Viral load in plasma

RNA was isolated from 140 µl of patient EDTA-plasma using column-based RNA isolation kit following the manufacturer’s instructions (Viral RNA mini kit, Qiagen). Isolated RNA was subjected to PUUV S RNA RT-qPCR analysis based on a previously described protocol,^63^ with TaqMan fast virus 1-step master mix (Thermo Scientific) using AriaMx instrumentation (Agilent).

### Flow cytometry analysis

PBMC samples were thawed in RPMI (Cytiva) complete media [L-glutamine (ThermoFisher Scientific), FCS (Sigma-Aldrich), penicillin/streptomycin (Cytiva)] with DNase (Roche) and counted. 3-4 million cells per sample were stained. Briefly, cells were incubated with LIVE/DEAD Fixable Green Dead Cell Stain Kit (ThermoFischer Scientific) -used as a viability marker- and fluorochrome-conjugated antibodies directed against surface markers (**Suppl Table 1**) for 20 min at room temperature in the dark followed by 2 washes with flow cytometry buffer (2 mM EDTA in PBS). Cells were then fixed and permeabilized using FACS Lysing solution and FACS Permeabilizing solution (BD Biosciences), and next incubated with intracellular staining antibodies (**Suppl Table 1**) for 30 min at 4°C in the dark. Cells were then washed and resuspended in flow cytometry buffer. Samples were acquired with a BD LSR Fortessa (BD Biosciences) flow cytometer. Flow cytometric analysis was performed using FlowJo version 10.7.2 (TreeStar, Ashland). To ensure unbiased manual gating, a blinded analysis was implemented, whereby all FCS3.0 files were renamed and coded by one person and blindly analyzed by another person. All samples were compensated electronically, and gatings were based on fluorescent-minus-one (FMO) or negative controls. After all gatings were performed, samples were decoded and data analysis was performed.

ILCs were defined as live (DCM^-^) CD1a^-^ CD14^-^ CD19^-^ CD34^-^ CD123^-^ BDCA2^-^ FcεR^-^ TCRα/β^-^ TCRγδ^-^ CD3^-^ CD45^+^ CD127^hi^. NK cells were defined as DCM^-^ CD1a^-^ CD14^-^ CD19^-^ CD34^-^ CD123^-^ BDCA2^-^ FcεR^-^ TCRα/β^-^ TCRγδ^-^CD3^-^ CD45^+^CD56^+^ CD127^lo/hi^. For a detailed gating strategy see **Suppl Fig 1.**

### Multiplex immunoassay

Plasma levels of IL-5, IL-6, IL-7, IL-10, IL-13, IL-15, IL-17A, IL-18, IL-23, IL-25, IL-33, IFN-γ, TNF, CCL20, CCL27, CCL28, GM-CSF, TSLP, and granzyme A were measured in plasma diluted 1:2, using a custom made Magnetic Luminex Screening assay (R&D Systems, Minneapolis) and analysed in a Magpix instrument (Luminex), according to manufacturer’s guidelines. For graphing purposes, values not calculated by the instrument’s software - because they were out of range in the lower standard range-were replaced by half of the value of the lowest detectable value for the given soluble factor measured.

### Statistical analysis

Statistical analyses were performed using GraphPad Prism software v.9.2 for MacOSX (GraphPad Software). Statistical differences between healthy controls and HFRS patients were analyzed with Kruskal-Wallis test followed by Dunn’s multiple comparison post hoc test. Paired comparisons between HFRS in the different disease stages (acute, early convalescence, and late convalescence) were performed using Wilcoxon test. p-values < 0.05 were considered statistically significant. Spearman’s rank correlation coefficient was used for assessing correlations. Spearman’s correlation matrixes were generated with R (v.4.1.1; R Core Team, 2020) using package corrplot (v.0.9). Principal component analysis (PCA) was performed in R (v.4.1.1) using packages Factoextra (v.1.0.7), FactoMineR (v.2.4), RColorBrewer (v.1.1-2), and ggplot2 (v.3.3.5). Data was normalised in R using the scale argument within the PCA function. Where data was missing, the values were imputed using package missMDA (v.1.18).

### Role of the funding sources

The funders of this study had no role in the study design, data collection, data analysis, data interpretation, or writing of the report.

## RESULTS

### Study design and patient characteristics

A total of 17 PUUV-infected hospitalized HFRS patients and 10 healthy controls were included in the study (**Table 1**). Peripheral blood samples were obtained at the acute (5-8 days after onset of symptoms), early convalescent (20-27 days after onset of symptoms), and late convalescent (180 or 360 days after onset of symptoms) phase of disease (**Fig 1a**). Hospitalized HFRS patients showed a typical clinical presentation during the acute phase, with thrombocytopenia, elevated C-reactive protein (CRP) and creatinine plasma levels, and viral load (**Table 1,** **Fig 1b, and Suppl Table 2**). For most of the patients all parameters normalized to levels within the normal range during the convalescent phase of disease (**Fig 1b**). The severity of the patients was assessed with a scoring system based on platelet counts, creatinine values, and mean arterial blood pressure values, as previously describe. ^61,62^ Two patients scored as severe, while all other scored as mild (**Suppl Table 2**).

**Figure 1.**
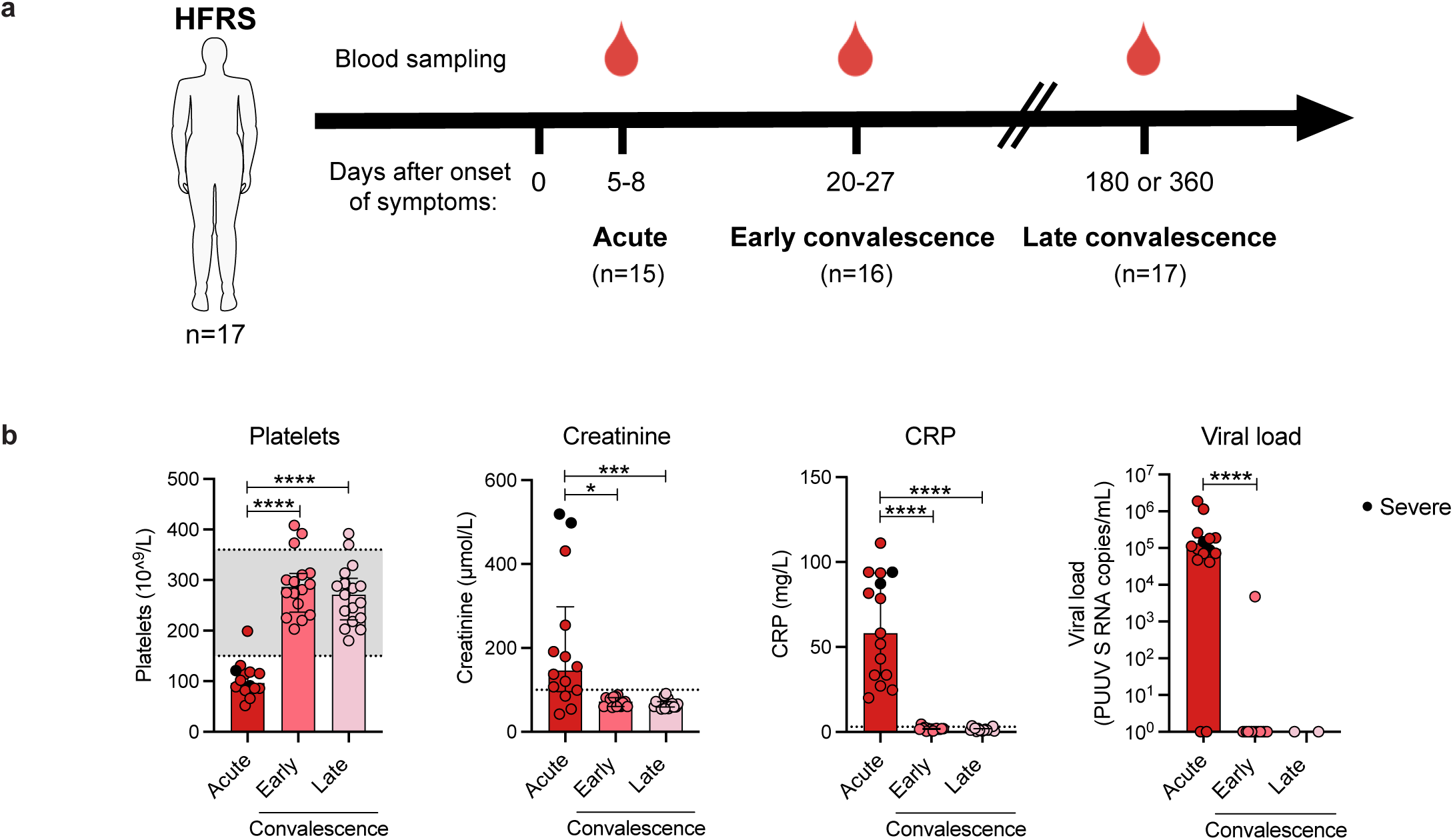
Cohort design and clinical characterization of PUUV-infected hemorrhagic fever with renal syndrome (HFRS) patients. **(a)** Schematic overview of peripheral blood sample collection in control subjects (n=10) and PUUV-infected HFRS patients (n=17) at acute (5 to 8 days), early convalescence (20 to 27 days), and late convalescence phase (180- or 360-days post onset of symptoms). **(b)** Levels of platelets (10^9^/L), creatinine (μmol/L), C-reactive protein (CRP, mg/L), and copy numbers of PUUV S RNA in plasma (10^4^/ml) in acute (n=14), early convalescence (n=15), and late convalescence (n=17) phase. Dotted lines indicate reference values in healthy adults. Graphs show data of individual subjects (circles) and the median (bars) ± interquartile range. Statistical significance was assessed using the Wilcoxon signed-rank test. Severe patients are indicated by a black circle. *p < 0.05; **p < 0.01; ***p < 0.001; ****p < 0.0001.

**Table 1.** Clinical and laboratory characteristics of donors.

| Characteristic | HFRS patients | Healthy controls |
| --- | --- | --- |
| n° of patients / controls | 17 | 10 |
| Age (years), median (IQR) | 35(28.5-51) | 47.5 (38.25-53.25) |
| Gender (female), n° (%) | 12 (71) | 2 (20) |
| Viral load at time of sampling (PUUV S RNA copies/mL), median (IQR) | 94,272 (47,300-187,975) | ND |
| Platelet count ($\times 10^9/L$), median (IQR) | 97 (85-119) | ND |
| CRP (mg/L), median (IQR) | 58.2 (33.4-93.5) | ND |
| Creatinine ($\mu\text{mol/L}$), median (IQR) | 146 (96-298) | ND |
| Days after symptoms onset, median (IQR) | 6 (6-8) | NA |
| Days hospitalized, median (IQR) | 5 (4 - 6.5) | NA |
| Hematocrit (L/L) median (IQR) | 0.37 (0.37-0.41) | ND |
| PCR positivity CMV, n° (%) | 0 (0) | ND |
| PCR positivity EBV, n° (%) | 1 (6) | ND |

NA: not applicable; ND: not determined; IQR: interquartile range; CRP: C-reactive protein; CMV: cytomegalovirus; EBV: Epstein-Barr virus
Platelet count; normal range $150\text{--}360 \times 10^9/L$ .
Plasma CRP; reference $<3 \text{ mg/L}$ .
Plasma creatinine; reference $<90 \mu\text{mol/L}$ for women, $<100 \mu\text{mol/L}$ for men.

### HFRS patients present a strong inflammatory response during the acute phase of disease

Hantavirus disease is characterized by strong systemic inflammatory responses.^37,40–43,64–67^ Using a multiplex immunoassay, we assessed the plasma levels of 19 cytokines in HFRS patients during the acute and convalescent phase of disease. Except for interleukin (IL)-5, IL-7, and IL-17A (**Suppl Fig 1a**), we observed differences in levels of all proteins between acute and later stages of HFRS (**Fig 2a** **and** **b**). As previously reported,^37,40–43,64–67^ the plasma levels of tumor necrosis factor (TNF), IL-6, granulocyte-macrophage colony-stimulating factor (GM-CSF), IL-10, interferon gamma (IFN-γ), IL-15, IL-18, and granzyme A (GrzA) were all significantly higher in acute HFRS as compared to the convalescent phases (**Fig 2b**). Further, we observed significantly higher levels of the type 2-associated cytokines IL-13, IL-25 (also called IL-17E), IL-33, and thymic stromal lymphopoietin (TSLP), related to ILC2 and T helper 2 cell activity and involved in functions such as tissue repair.^68,69^ We also observed significantly higher levels of the type 3-associated cytokine IL-23 in the acute phase of HFRS, involved in the activation of immune cells such as ILC3 and T helper 17 cells which have a role in protection from tissue damage (**Fig 2b**).^5,70^ Moreover, we observed that levels of the chemokines CCL20 and CCL27 were significantly higher in acute samples, while the level of CCL28 was significantly decreased (**Fig 2b**).

**Figure 2.**
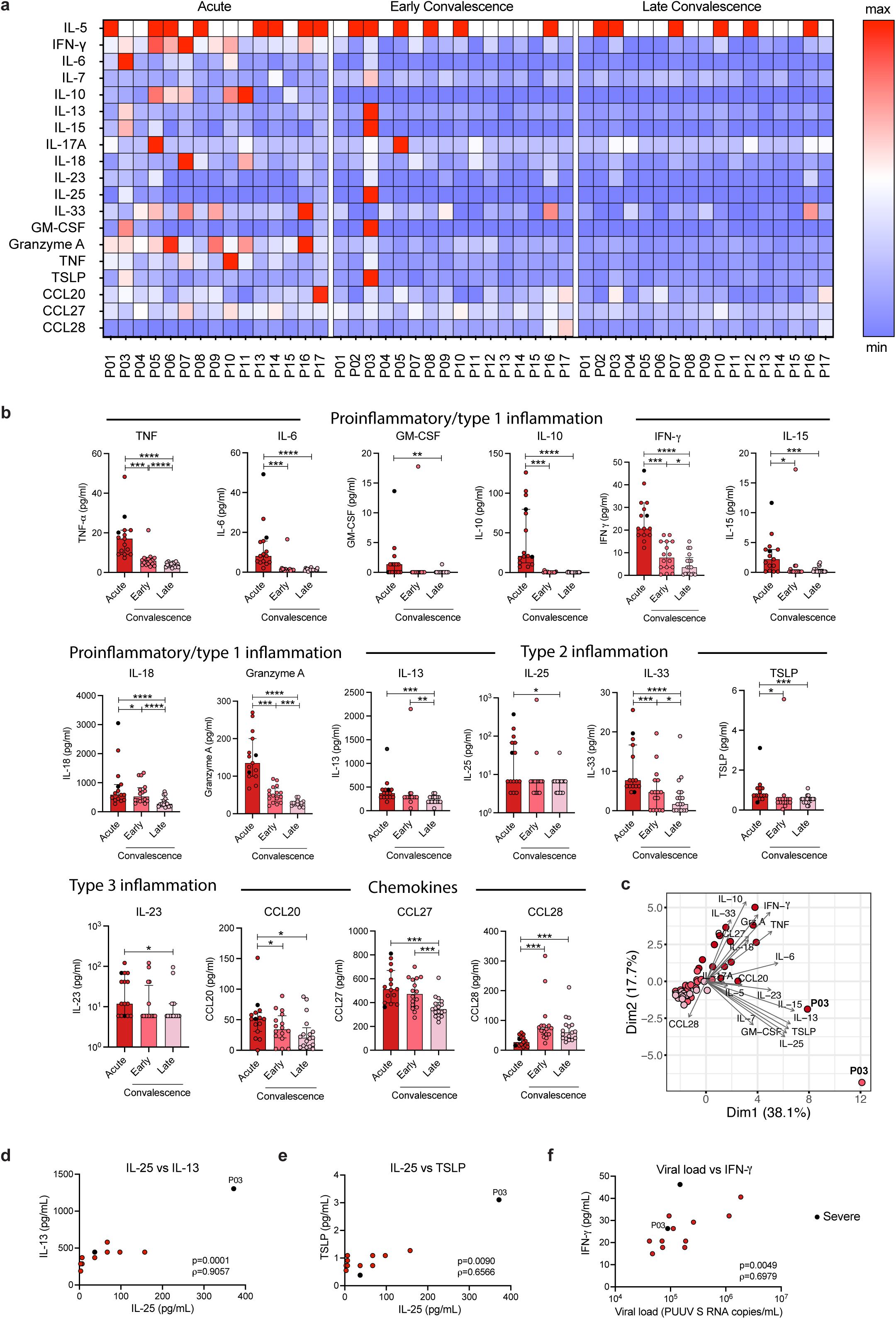
Altered levels of soluble factors in peripheral blood of HFRS patients. **(a)** Heatmap displaying the normalized plasma concentration of each soluble factor in each HFRS patient at acute, early convalescent, and late convalescent phase, as measured by multiplex immunoassay. Colors depict high (red) or low (blue) concentration. Values were normalized dividing each one by the highest value obtained for the given soluble factor. **(b)** Level of soluble factors in plasma of HFRS patients in acute (n=15), early convalescence (n=16), and late convalescence (n=17) phase. **(c)** Principal component analysis (PCA) displaying the distribution of HFRS patients according to the plasma level of soluble factors. **(d-f)** Spearman rank correlation between plasma levels of (**d)** IL-25 and IL-13, (**e**) IL-25 and TSLP, and (**f)** IFN-γ and viral load in acute HFRS patients (n=15). Abbreviations: IFN-γ: interferon gamma; IL: interleukin; TNF: tumor necrosis factor alpha; TSLP: Thymic stromal lymphopoietin; GM-CSF: granulocyte-macrophage colony-stimulating factor; CCL: chemokine ligand. Graphs show data of individual subjects (circles) and the median (bars) ± interquartile range. Statistical significance was assessed using the Wilcoxon signed-rank test. Severe patients are indicated by a black circle and patient 3 is labeled as P03. *p < 0.05; **p < 0.01; ***p < 0.001; ****p < 0.0001.

Principal component analysis (PCA) showed that samples from the acute phase separated from samples from the early and late convalescent phase. This separation was mainly driven by IL-10, GrzA, IFN-γ, TNF, IL-18, CCL27 and IL-33 (**Fig 2c**). Interestingly, patient 3, one of the two most severely ill patients in the cohort, deviated from the rest of the patients, both in the acute and early convalescent phase (**Fig 2c**). This patient showed the highest levels of several soluble factors and presented higher levels of many of them in the early convalescent phase than in the acute phase (**Fig 2a**), suggesting a longer than usual acute phase.

Additionally, we observed significant positive correlations between the type 2- associated cytokines IL-25 and IL-13 and between IL-25 and TSLP during the acute phase of HFRS (**Suppl Fig 1b** and **Fig 2d** **and** **e**). Further, out of the 15 acute HFRS patients, 13 were positive for PUUV S RNA in blood and a positive correlation was seen as well between viral load and IFN-γ (**Suppl Fig 1b** and **Fig 2f**).

Altogether, these results showed that PUUV-infected HFRS patients display a strong inflammatory response, including elevated levels of 16 cytokines many of which are known to be produced by or involved in the activation of ILCs and NK cells.

### Peripheral NK cells are activated but decreased in frequency during acute HFRS

We next characterized the ILC and NK cell compartments in PBMCs from HFRS patients. For the identification and analysis of ILCs and NK cells, we used 18-parameter flow cytometry and a modification of a well-established gating strategy (**Suppl Fig 2**).^71^

We observed decreased frequencies of total CD56^+^ NK cells in peripheral blood in the acute and early convalescent phase of disease, which normalized in late convalescence (**Fig 3a**). There was a decreased frequency of CD56^dim^ NK cells, with a concomitant increase of the smaller population of CD56^bright^ NK cells, during the acute phase of HFRS (**Fig 3b** **and** **c**). High frequencies of CD69^+^ and HLA-DR^+^ total CD56^+^ NK cells were detected in the acute phase of HFRS, indicating NK cell activation (**Fig 3d**). As expected from the general NK cell activation, frequencies of NKp44^+^ and NKG2A^+^ NK cells were also increased in the acute phase of HFRS (**Fig 3d**). Furthermore, the frequency of Ki-67^+^ NK cells was increased, showing that the NK cells proliferated during acute HFRS (**Fig 3d**). Additionally, when analyzing for homing receptors, we observed a decreased frequency of α4β7^+^ NK cells and an increased frequency of CCR6^+^ and CCR10^+^ NK cells in the acute phase of HFRS (**Fig 3d**), suggesting an effect on NK cell migration. The frequency of CD45RA^+^ and CD161^+^ NK cells was decreased in HFRS patients as compared to healthy controls, with a tendency to recovery in the convalescent phase of HFRS (**Fig 3d**). Analysis of surface markers in the CD56^bright^ and CD56^dim^ NK cell subsets showed similar results as observed for total NK cells (**Suppl Fig 3**). A PCA revealed a separation of acute HFRS from convalescent HFRS patients and healthy controls based on the level of expression of the different analyzed surface markers in total NK cells (**Fig 3e**).

**Figure 3.**
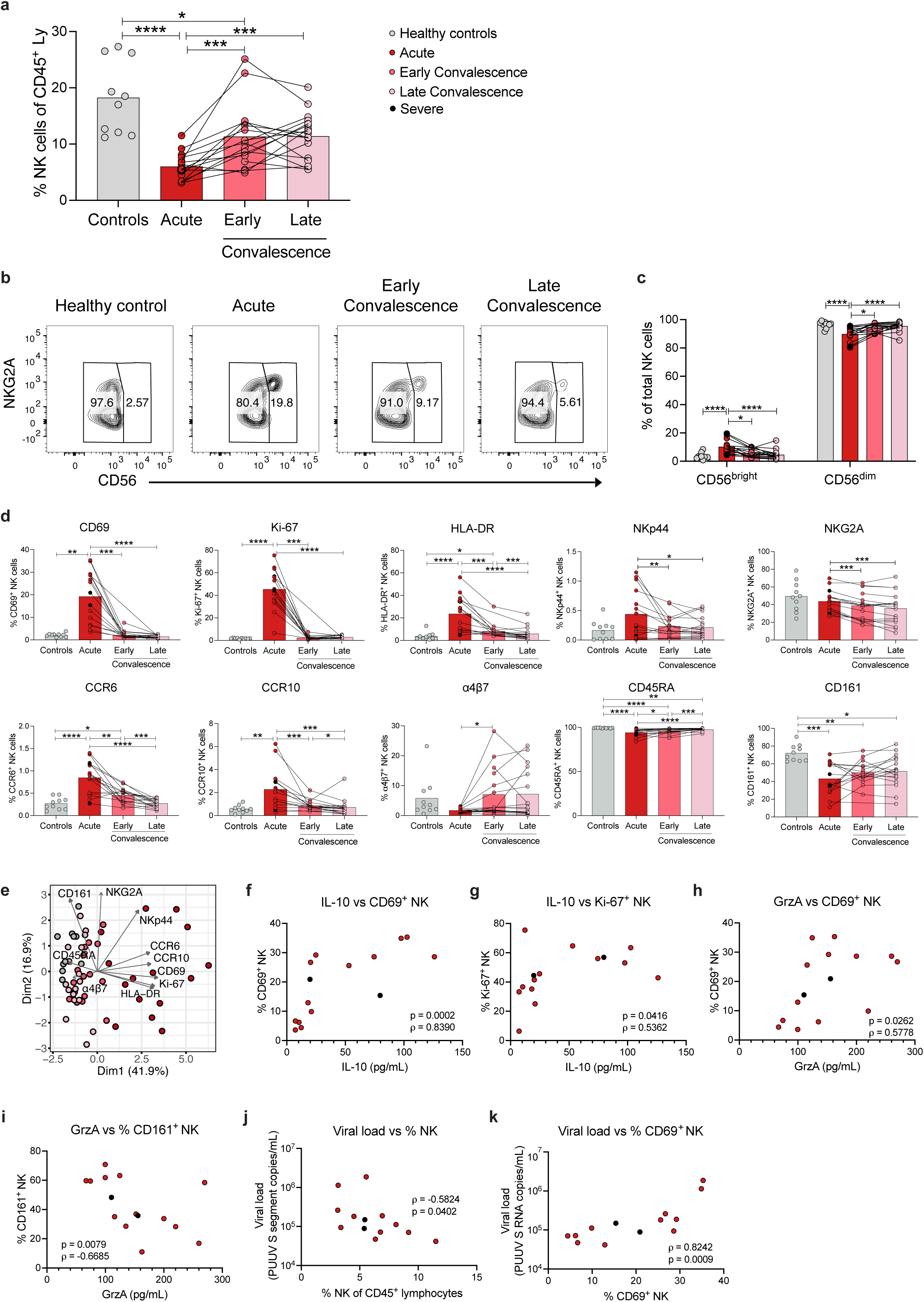
NK cells are activated, proliferative and decreased in frequency in peripheral blood of HFRS patients. **(a)** Percentage of NK cells out of CD45^+^ lymphocytes (Ly) in control donors (n=10) and HFRS patients during the acute (n=15), early convalescence (n=16), and late convalescence (n=17) phase. **(b)** Representative flow cytometry plots showing percentage of CD56^bright^ and CD56^dim^ NK cells (gated as CD3^-^CD56^+^ cells as in Fig S2) in a control donor and in an HFRS patient. (**c)** Percentage of CD56^bright^ and CD56^dim^ NK cells of CD3^-^CD56^+^ cells in control donors (n=10) and HFRS patients during the acute (n=15), early convalescence (n=16), and late convalescence (n=17) phase. **(d)** Percentage of CD69^+^, Ki-67^+^, HLA-DR^+^, NKp44^+^, NKG2A^+^, CCR6^+^, CCR10^+^, α4β7^+^, CD45RA^+^, and CD161^+^ NK cells of total CD3^-^CD56^+^ cells in control donors (n=10) and HFRS patients during the acute (n=15), early convalescence (n=16), and late convalescence (n=17) phase. **(e)** Principal component analysis (PCA) of total CD3^-^CD56^+^ NK cells in healthy controls and HFRS patients displaying the contribution of NK cell surface markers indicated in **(d)**. Each dot represents one donor. **(f-k)** Spearman rank correlation between plasma IL-10 levels and the percentage of (**f)** CD69^+^ NK cells and **g.** Ki-67^+^ NK cells, between plasma granzyme A (GrzA) levels and the percentage of **(h**) CD69^+^ NK cells and (**i)** CD161^+^ NK cells, and between plasma viral load (n=13; PUUV S RNA copies/mL) and the percentage of (**j)** NK cells out of CD45^+^ lymphocytes and (**k)** CD69^+^ NK cells in acute HFRS patients. Bar graphs are shown as mean and lines connect paired samples from the same patient. Statistical significance was assessed using the Wilcoxon signed-rank test to compare groups of HFRS patients, and the Kruskal-Wallis test followed by Dunn’s multiple comparisons test to compare healthy controls with groups of HFRS patients. Severe patients are indicated by black circles. *p < 0.05; **p < 0.01; ***p < 0.001; ****p < 0.0001.

Next, we examined correlations between frequencies of NK cells and both soluble plasma proteins and clinical parameters (**Suppl Fig 4a-d**). IL-10 plasma levels positively correlated with the frequencies of both activated (CD69^+^) and proliferating (Ki-67^+^) NK cells (**Fig 3f** **and** **g**). Granzyme A levels correlated positively with the frequency of CD69^+^ NK cells (**Fig 3h**), and negatively with the frequency of CD161^+^ NK cells (**Fig 3i**). Interestingly, the frequency of activated (CD69^+^) NK cells correlated positively with viral load while the frequency of total NK cells showed a negative correlation with viral load during the acute phase of HFRS (**Fig 3j** **and** **k**).

### Peripheral ILCs are activated and proliferate during acute HFRS

We next characterized the peripheral ILC responses in the HFRS patients. No significant difference in total ILC frequency was observed between the patients and the healthy controls (**Fig 4a**). Interestingly, as for NK cells, we observed a negative correlation between viral load and the frequency of ILCs in acute HFRS, showing reduced frequencies of peripheral ILCs in patients with higher viral loads (**Fig 4b**). Furthermore, we observed increased frequencies of activated (CD69^+^) and proliferating (Ki-67^+^) ILCs during the acute phase of HFRS (**Suppl Fig 5a**), while no differences were observed in the frequencies of NKp44, HLA-DR, and CD45RA-expressing ILCs (**Suppl Fig 5a**). When assessing expression of homing markers, we found a decreased frequency of α4β7^+^ ILCs in acute HFRS, but no significant difference in frequencies of ILCs expressing the chemokine receptors CCR6 and CCR10 (**Suppl Fig 5a**).

**Figure 4.**
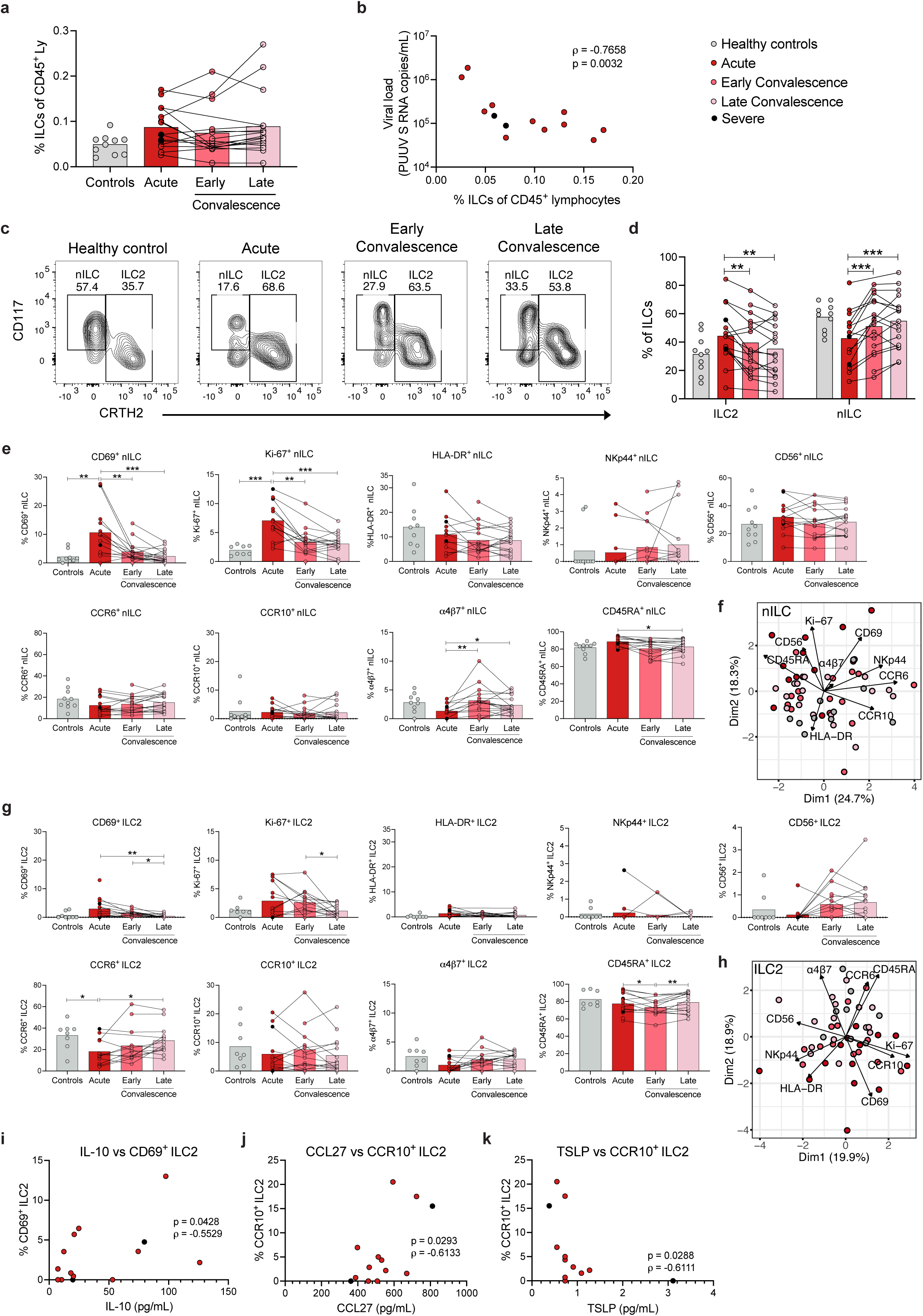
ILCs display an activated and proliferative profile in peripheral blood of HFRS patients. **(a)** Percentage of ILCs (gated as Lin^-^CD3^-^CD127^hi^ as in Fig S2) out of CD45^+^ lymphocytes (Ly) in control donors (n=10) and HFRS patients during the acute (n=15), early convalescence (n=16), and late convalescent (n=17) phase. **(b)** Spearman rank correlation between plasma viral load (PUUV S RNA copies/mL) and the percentage of ILCs out of CD45^+^ lymphocytes in acute HFRS patients (n=15). **(c)** Representative flow cytometry plots showing percentage of ILC2 and naïve ILC (nILC) in a control donor and in an HFRS patient (gated on ILCs as in Fig S2). (**d)** Percentage of ILC2 and nILC in control donors (n=10) and HFRS patients during the acute (n=15), early convalescence (n=16), and late convalescence (n=17) phase. **(e)** Percentage of CD69^+^, Ki-67^+^, HLA-DR^+^, NKp44^+^, CD56^+^, CCR6^+^, CCR10^+^, α4β7^+^, and CD45RA^+^ nILC in control donors (n=10) and HFRS patients during the acute (n=15), early convalescence (n=16), and late convalescence (n=17) phase. **(f)** Principal component analysis (PCA) of nILC in control donors and HFRS patients displaying the contribution of nILC surface markers indicated in **(e)**. **(g)** Percentage of CD69^+^, Ki-67^+^, HLA-DR^+^, NKp44^+^, CD56^+^, CCR6^+^, CCR10^+^, α4β7^+^, and CD45RA^+^ ILC2 in control donors (n=10) and HFRS patients during the acute (n=15), early convalescence (n=16), and late convalescence (n=17) phase. **(h)** PCA of ILC2 in control donors and HFRS patients displaying the contribution of ILC2 surface markers indicated in **(g)**. **(i-k)** Spearman rank correlation between (**i)** plasma IL-10 levels and the percentage of CD69^+^ ILC2, **(j)** plasma CCL27 levels and the percentage of CCR10^+^ ILC2, and (**k)** plasma TSLP levels and the percentage of CCR10^+^ ILC2 in acute HFRS patients (n=15). Bar graphs are shown as mean and lines connect paired samples from the same patient. Statistical significance was assessed using the Wilcoxon signed-rank test to compare groups of HFRS patients, and the Kruskal-Wallis test followed by Dunn’s multiple comparisons test to compare healthy controls with groups of HFRS patients. Severe patients are indicated by a black circle. Patients with low cell numbers (fewer than 20 events) in the corresponding gate were removed from the analysis. *p < 0.05; **p < 0.01; ***p < 0.001; ****p < 0.0001.

Next, we explored specific ILC subsets. CD117^neg^ ILCs in peripheral blood have been shown to make up a heterogenous population with yet undefined functions.^2^ We therefore decided to focus our analysis on the more well-defined nILC and ILC2. The composition of these ILC subsets changed over time in the HFRS patients (**Fig 4c**). We observed an increase in ILC2 frequency and a decreased frequency of naïve ILC (nILC) in the acute phase of HFRS (**Fig 4c** **and** **d**).

### Peripheral c-Kit^lo^ ILC2 are increased in frequency during HFRS

Next, we characterized the phenotype of the ILC subsets. Increased frequencies of activated (CD69^+^) and proliferating (Ki-67^+^) nILC were observed in acute HFRS (**Fig 4e**). Moreover, similar to NK cells (**Fig 3d**), a decreased frequency of α4β7^+^ nILC was observed in acute HFRS (**Fig 4e**). No differences were observed in the frequencies of nILCs expressing HLA-DR, NKp44, CCR6, and CCR10 in HFRS patients as compared to healthy controls (**Fig 4e**). PCA based on the frequency of expression of surface markers in nILC showed a separation between the acute HFRS patients and healthy controls (**Fig 4f**).

The ILC2 population showed a similar phenotypic pattern as the nILC, with increased frequency of activated (CD69^+^) and proliferating (Ki-67^+^) cells during acute HFRS (**Fig 4g**). Additionally, a significantly decreased frequency of CCR6^+^ ILC2 was observed in the acute phase of disease (**Fig 4g**), while no differences were observed in the frequencies of ILC2 expressing NKp44, HLA-DR, CCR10, and α4β7 in HFRS as compared to healthy controls (**Fig 4g**). In line with these findings, a PCA based on the frequency of expression of surface markers on ILC2s showed no clear separation between the acute and convalescent HFRS, but a separation of the acute and control samples was observed (**Fig 4h**). Further, when analyzing for possible correlations to soluble proteins (**Suppl Fig 6**), we observed a positive correlation between the plasma levels of IL-10 and the frequency of CD69^+^ ILC2 (**Fig 4i**) and between plasma levels of the CCR10 ligand CCL27 and CCR10^+^ ILC2 (**Fig 4j**), as well as a negative correlation between plasma levels of TSLP and frequency of CCR10^+^ ILC2 in acute HFRS patients (**Fig 4k**). The first two correlations were also observed for total ILCs, as well as a positive correlation between plasma levels of IL-7 and Ki-67^+^ ILC (**Suppl Fig 5b)**.

Having observed an increased frequency of ILC2 (**Fig 4d**), a decreased frequency of CCR6^+^ ILC2 (**Fig 4g**), and increased plasma levels of type 2 cytokines in acute HFRS (**Fig 2**), we next assessed whether there were changes in the ILC2 subsets in HFRS patients. Indeed, we observed increased frequencies of c-Kit^lo^ ILC2 and, concomitantly, decreased frequencies of c-Kit^hi^ ILC2 during the acute phase of HFRS, as compared to convalescent HFRS patients and healthy controls (**Fig 5a** **and** **b**). Moreover, c-Kit^hi^ ILC2 showed higher frequency of CCR6 expression than c-Kit^lo^ ILC2 both in patients and healthy controls (**Fig 5c**) and, aligning with the relative depletion of CCR6^+^ ILC2 in acute HFRS, (**Fig 4g**) we observed a lower frequency of CCR6^+^ c-Kit^hi^ ILC2 in the acute phase of HFRS as compared to the convalescent phase (**Fig 5c**).

**Figure 5.**
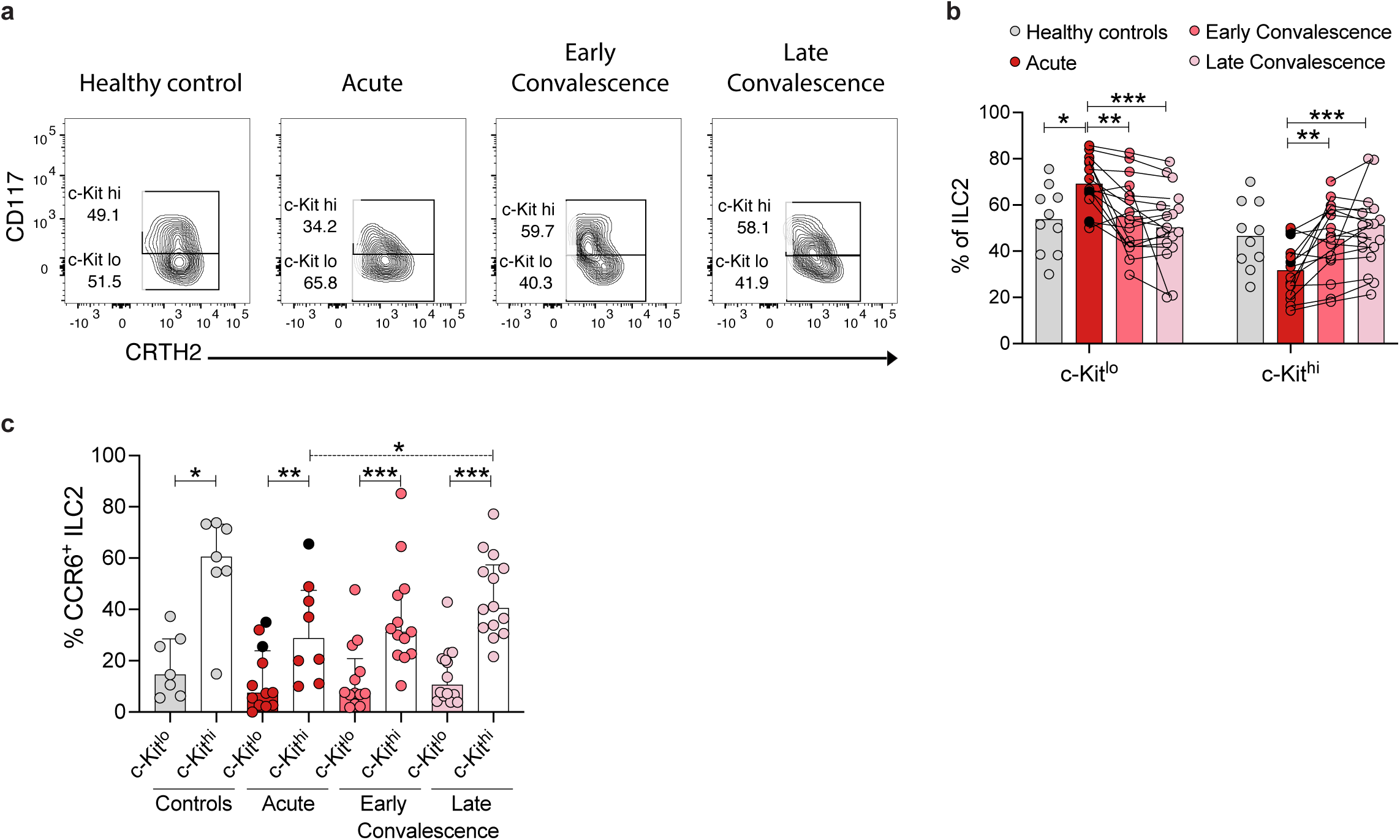
Increased ILC2 c-Kit^lo^ frequency in peripheral blood of acute HFRS patients. **(a)** Representative flow cytometry plots showing percentages of c-Kit^lo^ and c-Kit^hi^ ILC2 in a control donor and in an HFRS patient (gated on total ILC2 as in Fig S2). **(b)** Percentage of c-Kit^lo^ and c-Kit^hi^ ILC2 in control donors (n=10) and HFRS patients during the acute (n=15), early convalescence (n=16), and late convalescence (n=17) phase. **(c)** Percentage of CCR6^+^ c-Kit^lo^ and CCR6^+^ c-Kit^hi^ ILC2 in control donors (n=10) and HFRS patients during the acute (n=15), early convalescence (n=16), and late convalescence (n=17) phase. Bar graphs are shown as mean and lines connect paired samples from the same patient. Statistical significance was assessed using the Wilcoxon signed-rank test to compare groups of HFRS patients, and the Kruskal-Wallis test followed by Dunn’s multiple comparisons test to compare healthy controls with groups of HFRS patients. Severe patients are indicated by a black circle. Patients with low cell numbers (fewer than 20 events) in the corresponding gate were removed from the analysis. *p < 0.05; **p < 0.01; ***p < 0.001.

## DISCUSSION

Here we provide a detailed characterization of total ILCs, including both NK cells and non-NK ILCs, in blood samples from PUUV-infected HFRS patients. We reveal that total ILCs are activated and show an altered composition correlating to viral load.

Whereas NK cells have been extensively described in several human viral infections ^8–10^ including hantavirus infections, ^46–49,72,73^ ILCs remain understudied in human viral infections. Recent studies reveal dysregulated frequencies and phenotypes of ILCs in HIV-1,^21,22^ SARS-CoV2,^23–25^ rhinovirus,^74^ and respiratory syncytial virus (RSV) infection.^26^ Here we characterized ILCs and NK cell responses in the context of human hantavirus infection.

HFRS is characterized by a strong general immune activation, including hyperinflammation. ^30,75^ Some typical laboratory features are leukocytosis, thrombocytopenia, high CRP and hematocrit levels, and due to acute kidney injury, increased serum creatinine, proteinuria, and haematuria.^28,31,76–78^ Viral load peaks between the first 3 to 5 days during the febrile phase of disease, and viral load is normally not detectable in the convalescent phase of disease ^31,32,79,80^. In line with this, the HFRS patients in our study presented with typical laboratory findings together with a strong inflammatory response in the acute phase of disease.^37,40–43,64–67^ We observed an increase of several type 2-associated cytokines such as IL-13, IL-25, IL-33, and TSLP in acute PUUV-infected HFRS patients. The alarmins IL-25, IL-33, and TSLP are known activators of ILC2, which upon activation can secrete IL-13.^15,81^ These alarmins are upregulated upon infection or damage mainly of epithelial, endothelial, and stromal cells.^82–84^ Given that the main target cell of hantaviruses are endothelial cells,^30^ it is likely that hantavirus induce the secretion of the mentioned alarmins by these cells in HFRS patients. Interestingly, IL-33 has previously been found to be also elevated in plasma of Hantaan virus-infected HFRS patients and to positively correlate with disease severity.^85^

As recently shown by Resman Rus and colleagues,^48^ we observed transiently reduced NK cell frequencies in peripheral blood of PUUV-infected HFRS patients, returning to normal levels during convalescence. Moreover, as earlier shown,^47^ we observed that remaining circulatory NK cells were highly proliferating, with approximately half of them expressing Ki-67. During the acute phase of HFRS, NK cells also showed increased expression of several activation markers and altered expression of chemokine receptors associated with migration to tissues, such as lung (CCR6) and intestine (α4β7).^86–89^ This suggests migration of NK cells to tissues, which could be the cause of the observed decreased frequencies of NK cells in peripheral blood during acute HFRS. Interestingly, viral load correlated negatively with the frequency of NK cells in peripheral blood but positively with the frequency of activated NK cells, suggesting that active viral replication impacts, in an unknown manner, the activation of NK cells during HFRS. We have previously shown that hantaviruses have strong anti-apoptotic properties that potentially protect infected cells from cytotoxic lymphocyte mediated killing and may trigger NK cell mediated bystander killing of uninfected cells.^72,73,90,91^ Whether this holds true *in vivo* remains to be investigated, but the strong activation of NK cells we observe in HFRS patients could be a sign of continuous attempts of NK cells to kill hantavirus-infected cells. Of further interest, we observed that levels of IFN-γ also positively correlated with viral load in HFRS patients. IFN-γ is a cytokine with strong antiviral effects produced by a wide array of innate and adaptive lymphocytes, including ILCs, NK cells, NKT cells, and T cells.^92–95^ The main targets of hantavirus infection, such as endothelial cells and monocytes,^30,59^ are not known to express IFN-γ, so this correlation further suggests that active viral replication in infected cells indirectly leads to enhanced IFN-γ production by immune cells, such as activated NK cells.

We observed a decreased frequency of CD161^+^ NK cells in acute HFRS patients. The role of CD161 in NK cells is not fully understood, but it has been suggested to mark pro-inflammatory NK cells with a high ability to respond to innate cytokines.^96^ There are previous reports of modulation of CD161 expression in NK cells in other viral infections, such as a reduced CD161 expression in NK cells in acute hepatitis C virus infection which associated with enhanced viral clearance,^97^ reduced frequency of CD161^+^ NK cells in Chikungunya-infected patients,^98^ and depletion of CD161^+^ NK cells in cytomegalovirus infection.^96^ Paralleling the findings on NK cells, downregulation of CD161 expression on MAIT cells has been described in HFRS,^42^ as well as in HIV-infected individuals.^99^ Interestingly, we also observed a negative correlation between the plasma levels of granzyme A and the frequency of CD161^+^ NK cells in acute HFRS. Even though NK cells are not the sole source of granzyme A in plasma, this observation could suggest that the decrease in frequency of CD161^+^ NK cells - and thus increase of a CD161^-^ NK population - is related to higher NK cell degranulation and cytotoxicity for viral control in HFRS patients. Along this line, we also observed a positive correlation between activated NK cells and the levels of IL-10, a cytokine that has been reported to enhance the effector functions of NK cells.^100^

The frequency of ILCs is reduced in the circulation of acute HIV-infected individuals, and correlates negatively with viral load.^22^ In COVID-19 we also recently reported a decrease in peripheral ILC frequencies and numbers.^23^ In contrast, here we did not observe a significant change in the frequency of ILCs in HFRS as compared to healthy controls. Interestingly though, and in line with the previous report for HIV infection,^22^ frequencies of peripheral ILCs in acute HFRS showed a strong negative correlation with viral load. Furthermore, as described for moderate COVID-19 patients,^23^ acute HFRS patients presented increased frequencies of ILC2 with a concomitant reduction of nILC frequencies. In contrast to our findings in COVID-19 patients,^23^ we observed increased frequencies of Ki-67-expressing ILC2 and nILC in HFRS patients. Moreover, the decreased levels of α4β7^+^ nILCs in acute HFRS suggest that the reduced frequency of peripheral blood nILCs could be due to their migration to tissues ^4^. Alternatively, nILC might differentiate into mature ILC subsets, such as ILC2s, in the circulation. Of interest, acute HFRS patients presented increased frequencies of c-Kit^lo^ ILC2. Two ILC2 subsets have been defined, differing in their surface expression of c-Kit and their functionality. c-Kit^lo^ ILC2 are more mature and ILC2-lineage committed, while c-Kit^hi^ ILC2 show plasticity towards an ILC3 phenotype and functionality.^7^ Moreover, c-Kit^lo^ ILC2 express less CCR6 compared to c-Kit^hi^ ILC2.^7^ In line with this, acute HFRS patients showed decreased frequency of CCR6^+^ c-Kit^hi^ ILC2 as compared to convalescent patients and healthy controls. This suggests a skewing of c-Kit^hi^ ILC2 towards more ILC2-commited cells in HFRS, possibly explaining the increase in c-Kit^lo^ ILC2 frequencies in acute HFRS. Alternatively, this decrease in CCR6^+^ c-Kit^hi^ ILC2 levels could also be due to migration of these cells to lungs, where ILC2 have been shown to play an important role in lung tissue repair during influenza infection in mice.^15,16^

Here we have characterized peripheral ILCs in PUUV-infected HFRS patients. Future characterization of ILCs in tissue samples, such as lung and intestines, can add important knowledge regarding circulating ILC infiltration and local ILC responses.

In conclusion, this study provides the first comprehensive characterization of total circulating ILCs in hantavirus-infected patients. We report an overall activated and proliferating ILC profile in these patients, with a particular increased frequency of the ILC2 subset, and a skewing towards the ILC2-lineage committed c-Kit^lo^ ILC2 in acute HRFS. Additionally, we show that NK cells are reduced in frequencies and confirm that remaining circulating NK cells are highly activated and proliferating in acute HFRS. Moreover, we report a negative correlation between viral load and the frequencies of both NK cells and ILCs in acute HFRS, suggesting a potential influence of viral replication on these cells during the acute phase of hantavirus-caused disease.

## Data Availability

The authors declare that the data supporting the findings of this study are available from the corresponding authors upon reasonable request.

## CONTRIBUTORS

All authors read and approved the final version of the manuscript.

MG: data curation, funding acquisition, formal analysis, investigation, methodology, visualization, writing-original draft, writing-review & editing;

ACG: data curation, formal analysis, investigation, methodology, visualization, writing-review & editing;

JT: methodology;

KM: methodology;

AV: resources, funding acquisition;

SM: resources, funding acquisition;

JuM: resources, funding acquisition;

ASS: resources;

TS: resources; funding acquisition;

JeM: conceptualization, investigation, visualization, writing-review & editing, supervision;

JK: conceptualization, funding acquisition, investigation, visualization, writing-review & editing, supervision.

## DECLARATION OF INTERESTS

The authors have no competing financial interests or conflicts.

## ACKNOWLEDGMENTS

We thank the patients and volunteers who have contributed with clinical material to this study. We also thank Ms. Sanna Mäki for expert technical assistance. This study was supported by grants from the Swedish Research Council (projects K2015-56X-22774-01-3 and 2018-02646 to JK), Karolinska Institutet Research Foundation (project 2020-01469 to MG), the Academy of Finland (project 321809 to TS), Magnus Ehrnrooth Foundation (to AV), and Sigrid Jusélius Foundation (to AV and JuM), and the Competitive State Research Financing of the Responsibility Area of Tampere University Hospital (9AA050 and 9AB046 to JuM and 9AA052 to SM).

## Notes

### Competing Interest Statement

The authors have declared no competing interest.

### Funding Statement

This study was funded by grants from the Swedish Research Council (projects K2015-56X- 22774-01-3 and 2018-02646 to JK), Karolinska Institutet Research Foundation (project 2020-01469 to MG), the Academy of Finland (project 321809 to TS), Magnus Ehrnrooth Foundation (to AV), and Sigrid Juselius Foundation (to AV and JuM), and the Competitive State Research Financing of the Responsibility Area of Tampere University Hospital (9AA050 and 9AB046 to JuM and 9AA052 to SM).

### Author Declarations

The study was approved by the Ethics Committee of Tampere University Hospital (ethical permit nr. R04180), and the Swedish ethics review authority (etikprovningsmyndigheten, ethical permit nr. 2020-02604)

## REFERENCES

1. Sonnenberg GF, Hepworth MR. Functional interactions between innate lymphoid cells and adaptive immunity. Nat Rev Immunol 2019; 19: 599–613. doi:10.1038/s41577-019-0194-8

2. Mazzurana L, Czarnewski P, Jonsson V, Wigge L, Ringnér M, Williams TC, et al. Tissue-specific transcriptional imprinting and heterogeneity in human innate lymphoid cells revealed by full-length single-cell RNA-sequencing. Cell Res 2021; 31: 554–68. doi:10.1038/s41422-020-00445-x

3. Kokkinou E, Pandey RV, Mazzurana L, Gutierrez-Perez I, Tibbitt CA, Weigel W, et al. CD45RA + CD62L − ILCs in human tissues represent a quiescent local reservoir for the generation of differentiated ILCs. Sci Immunol 2022; 7: eabj8301. doi:10.1126/sciimmunol.abj8301

4. Lim AI, Li Y, Lopez-Lastra S, Stadhouders R, Paul F, Casrouge A, et al. Systemic Human ILC Precursors Provide a Substrate for Tissue ILC Differentiation. Cell 2017; 168: 1086–100. doi:10.1016/j.cell.2017.02.021

5. Vivier E, Artis D, Colonna M, Diefenbach A, Di Santo JP, Eberl G, et al. Innate Lymphoid Cells: 10 Years On. Cell 2018; 174: 1054–66. doi:10.1016/j.cell.2018.07.017

6. Spits H, Artis D, Colonna M, Diefenbach A, Di Santo JP, Eberl G, et al. Innate lymphoid cells-a proposal for uniform nomenclature. Nat Rev Immunol 2013; 13: 145–9. doi:10.1038/nri3365

7. Hochdörfer T, Winkler C, Pardali K, Mjösberg J. Expression of c-Kit discriminates between two functionally distinct subsets of human type 2 innate lymphoid cells. Eur J Immunol 2019; 49: 884–93. doi:10.1002/eji.201848006

8. Jost S, Altfeld M. Control of human viral infections by natural killer cells. Annu Rev Immunol 2013; 31: 163–94. doi:10.1146/annurev-immunol-032712-100001

9. Brandstadter JD, Yang Y. Natural killer cell responses to viral infection. J Innate Immun 2011; 3: 274–9. doi:10.1159/000324176

10. Lam VC, Lanier LL. NK cells in host responses to viral infections. Curr Opin Immunol 2017; 44: 43–51. doi:10.1016/j.coi.2016.11.003.

11. Mebius RE, Rennert P, Weissman IL. Developing lymph nodes collect CD4+CD3-LTβ+ cells that can differentiate to APC, NK cells, and follicular cells but not T or B cells. Immunity 1997; 7: 493–504. doi:10.1016/S1074-7613(00)80371-4

12. Spits H, Di Santo JP. The expanding family of innate lymphoid cells: Regulators and effectors of immunity and tissue remodeling. Nat Immunol 2011; 12: 21–7. doi:10.1038/ni.1962

13. Hildreth AD, O’sullivan TE. Tissue-resident innate and innate-like lymphocyte responses to viral infection. Viruses 2019; 11. doi:10.3390/v11030272

14. Stehle C, Hernández DC, Romagnani C. Innate lymphoid cells in lung infection and immunity. Immunol Rev 2018; 286: 102–19. doi:10.1111/imr.12712

15. Monticelli LA, Sonnenberg GF, Abt MC, Alenghat T, Ziegler CGK, Doering TA, et al. Innate lymphoid cells promote lung-tissue homeostasis after infection with influenza virus. Nat Immunol 2011; 12: 1045–54. doi:10.1038/ni.2131

16. Califano D, Furuya Y, Roberts S, Avram D, McKenzie ANJ, Metzger DW. IFN-γ increases susceptibility to influenza A infection through suppression of group II innate lymphoid cells. Mucosal Immunol 2018; 11: 209–19. doi:10.1038/mi.2017.41

17. Chang YJ, Kim HY, Albacker LA, Baumgarth N, McKenzie ANJ, Smith DE, et al. Innate lymphoid cells mediate influenza-induced airway hyper-reactivity independently of adaptive immunity. Nat Immunol 2011; 12: 631–8. doi:10.1038/ni.2045

18. Shim DH, Park YA, Kim MJ, Hong JY, Baek JY, Kim KW, et al. Pandemic influenza virus, pH1N1, induces asthmatic symptoms via activation of innate lymphoid cells. Pediatr Allergy Immunol 2015; 26: 780–8. doi:10.1111/pai.12462

19. Li BWS, de Bruijn MJW, Lukkes M, van Nimwegen M, Bergen IM, KleinJan A, et al. T cells and ILC2s are major effector cells in influenza-induced exacerbation of allergic airway inflammation in mice. Eur J Immunol 2019; 49: 144–56. doi:10.1002/eji.201747421

20. Stier MT, Bloodworth MH, Toki S, Newcomb DC, Goleniewska K, Boyd KL, et al. Respiratory syncytial virus infection activates IL-13–producing group 2 innate lymphoid cells through thymic stromal lymphopoietin. J Allergy Clin Immunol 2016; 138: 814–824.e11. doi:10.1016/j.jaci.2016.01.050

21. Krämer B, Goeser F, Lutz P, Glässner A, Boesecke C, Schwarze-Zander C, et al. Compartment-specific distribution of human intestinal innate lymphoid cells is altered in HIV patients under effective therapy. PLoS Pathog 2017; 13. doi:10.1371/journal.ppat.1006373

22. Kløverpris HN, Kazer SW, Mjösberg J, Mabuka JM, Wellmann A, Ndhlovu Z, et al. Innate Lymphoid Cells Are Depleted Irreversibly during Acute HIV-Infection in the Absence of Viral Suppression. Immunity 2016; 44: 391–405. doi:10.1016/j.immuni.2016.01.006

23. García M, Kokkinou E, Carrasco García A, Parrot T, Palma Medina LM, Maleki KT, et al. Innate lymphoid cell composition associates with COVID-19 disease severity. Clin Transl Immunol 2020; 9: e1224. doi:10.1002/cti2.1224

24. Gomez-Cadena A, Spehner L, Kroemer M, Khelil M Ben, Bouiller K, Verdeil G, et al. Severe COVID-19 patients exhibit an ILC2 NKG2D+ population in their impaired ILC compartment. Cell Mol Immunol 2021; 18: 484–6. doi:10.1038/s41423-020-00596-2

25. Gomes AM, Farias GB, Dias-Silva M, Laia J, Trombetta AC, Godinho-Santos A, et al. SARS-CoV2 pneumonia recovery is linked to expansion of Innate Lymphoid Cells type 2 expressing CCR10. Eur J Immunol 2021; 0: 1–8. doi:10.1002/eji.202149311

26. Vu LD, Siefker D, Jones TL, You D, Taylor R, DeVincenzo J, et al. Elevated levels of type 2 respiratory innate lymphoid cells in human infants with severe respiratory syncytial virus bronchiolitis. Am J Respir Crit Care Med 2019; 200: 1414–23. doi:10.1164/rccm.201812-2366OC

27. Kuhn JH, Adkins S, Agwanda BR, Al Kubrusli R, Alkhovsky S V., Amarasinghe GK, et al. 2021 Taxonomic update of phylum Negarnaviricota (Riboviria: Orthornavirae), including the large orders Bunyavirales and Mononegavirales. Arch Virol 2021; 166: 3513–66. doi:10.1007/s00705-021-05143-6

28. Jonsson CB, Figueiredo LTM, Vapalahti O. A global perspective on hantavirus ecology, epidemiology, and disease. Clin Microbiol Rev 2010; 23: 412–41.

29. Sabino-Santos Jr G, Gonçalves F, Martins RB, Gagliardi TB, Souza WM De, Muylaert RL, et al. Natural infection of Neotropical bats with hantavirus in Brazil. Sci Rep 2018; 8: 1–8. doi:10.1038/s41598-018-27442-w

30. Vaheri A, Strandin T, Hepojoki J, Sironen T, Henttonen H, Mäkelä S, et al. Uncovering the mysteries of hantavirus infections. Nat Rev Microbiol 2013; 11: 539–50. doi:10.1038/nrmicro3066

31. Avšič-Županc T, Saksida A, Korva M. Hantavirus infections. Clin Microbiol Infect 2019; 21: e6–16. doi:10.1111/1469-0691.12291

32. Klingström J, Smed-Sörensen A, Maleki KT, Solà-Riera C, Ahlm C, Björkström NK, et al. Innate and adaptive immune responses against human Puumala virus infection: immunopathogenesis and suggestions for novel treatment strategies for severe hantavirus-associated syndromes. J Intern Med 2019; 285: 510–23. doi:10.1111/joim.12876

33. Vaheri A, Henttonen H, Mustonen J. Hantavirus research in finland: Highlights and perspectives. Viruses 2021; 13: 1452. doi:10.3390/v13081452

34. Jiang H, Du H, Wang LM, Wang PZ, Bai XF. Hemorrhagic Fever with Renal Syndrome: Pathogenesis and Clinical Picture. Front Cell Infect Microbiol 2016; 6: 1. doi:10.3389/fcimb.2016.00178

35. Liu R, Ma H, Shu J, Zhang Q, Han M, Liu Z, et al. Vaccines and Therapeutics Against Hantaviruses. Front Microbiol 2020; 10. doi:10.3389/fmicb.2019.02989

36. Terajima M, Ennis FA. T cells and pathogenesis of hantavirus cardiopulmonary syndrome and hemorrhagic fever with renal syndrome. Viruses 2011; 3: 1059–73. doi:10.3390/v3071059

37. Sadeghi M, Eckerle I, Daniel V, Burkhardt U, Opelz G, Schnitzler P. Cytokine expression during early and late phase of acute Puumala hantavirus infection. BMC Immunol 2011; 12. doi:10.1186/1471-2172-12-65

38. Linderholm M, Ahlm C, Settergren B, Waage A, Tärnvik A. Elevated plasma levels of tumor necrosis factor (TNF)-α, soluble TNF receptors, interleukin (IL)-6, and IL-10 in patients with hemorrhagic fever with renal syndrome. J Infect Dis 1996; 173: 38–43. doi:10.1093/infdis/173.1.38

39. Borges AA, Campos GM, Moreli ML, Moro Souza RL, Saggioro FP, Figueiredo GG, et al. Role of mixed Th1 and Th2 serum cytokines on pathogenesis and prognosis of hantavirus pulmonary syndrome. Microbes Infect 2008; 10: 1150–7. doi:10.1016/j.micinf.2008.06.006

40. Saksida A, Wraber B, Avšič-Županc T. Serum levels of inflammatory and regulatory cytokines in patients with hemorrhagic fever with renal syndrome. BMC Infect Dis 2011; 11. doi:10.1186/1471-2334-11-142

41. Morzunov SP, Khaiboullina SF, St Jeor S, Rizvanov AA, Lombardi VC. Multiplex Analysis of Serum Cytokines in Humans with Hantavirus Pulmonary Syndrome. Front Immunol 2015; 6: 432. doi:10.3389/fimmu.2015.00432

42. Maleki KT, Tauriainen J, García M, Kerkman PF, Christ W, Dias J, et al. MAIT cell activation is associated with disease severity markers in acute hantavirus infection. Cell Reports Med 2021; 2: 100220. doi:10.1016/j.xcrm.2021.100220

43. Maleki KT, García M, Iglesias A, Alonso D, Ciancaglini M, Hammar U, et al. Serum Markers Associated with Severity and Outcome of Hantavirus Pulmonary Syndrome. J Infect Dis 2019; 219: 1832–40. doi:10.1093/infdis/jiz005

44. Srikiatkhachorn A, Spiropoulou CF. Vascular events in viral hemorrhagic fevers: A comparative study of dengue and hantaviruses. Cell Tissue Res 2014; 355: 621–33. doi:10.1007/s00441-014-1841-9

45. Hepojoki J, Vaheri A, Strandin T. The fundamental role of endothelial cells in hantavirus pathogenesis. Front Microbiol 2014; 5: 1–7. doi:10.3389/fmicb.2014.00727

46. Linderholm M, Bjermer L, Juto P, Roos G, Sandström T, Settergren B, et al. Local host response in the lower respiratory tract in nephropathia epidemica. Scand J Infect Dis 1993; 25: 639–46. doi:10.3109/00365549309008554

47. Björkström NK, Lindgren T, Stoltz M, Fauriat C, Braun M, Evander M, et al. Rapid expansion and long-term persistence of elevated NK cell numbers in humans infected with hantavirus. J Exp Med 2011; 208: 13–21. doi:10.1084/jem.20100762

48. Resman Rus K, Kopitar AN, Korva M, Ihan A, Petrovec M, Avšič-Županc T. Comparison of Lymphocyte Populations in Patients With Dobrava or Puumala orthohantavirus Infection. Front Cell Infect Microbiol 2020; 10: 566149. doi:10.3389/fcimb.2020.566149

49. Vietzen H, Hartenberger S, Aberle SW, Puchhammer-Stöckl E. Dissection of the NKG2C NK cell response against Puumala Orthohantavirus. Holbrook MR, editor. PLoS Negl Trop Dis 2021; 15: e0010006. doi:10.1371/JOURNAL.PNTD.0010006

50. García M, Iglesias A, Landoni VI, Bellomo C, Bruno A, Córdoba MT, et al. Massive plasmablast response elicited in the acute phase of hantavirus pulmonary syndrome. Immunology 2017; 151: 122–35. doi:10.1111/imm.12713

51. Rasmuson J, Pourazar J, Mohamed N, Lejon K, Evander M, Blomberg A, et al. Cytotoxic immune responses in the lungs correlate to disease severity in patients with hantavirus infection. Eur J Clin Microbiol Infect Dis 2016; 35: 713–21. doi:10.1007/s10096-016-2592-1

52. Iglesias AA, Períolo N, Bellomo CM, Lewis LC, Olivera CP, Anselmo CR, et al. Delayed viral clearance despite high number of activated T cells during the acute phase in Argentinean patients with hantavirus pulmonary syndrome. eBioMedicine 2022; 75: 103765. doi:10.1016/j.ebiom.2021.103765

53. Kerkman PF, Dernstedt A, Tadala L, Mittler E, Dannborg M, Sundling C, et al. Generation of plasma cells and CD27−IgD− B cells during hantavirus infection is associated with distinct pathological findings. Clin Transl Immunol 2021; 10: e1313. doi:10.1002/cti2.1313

54. Hepojoki J, Cabrera LE, Hepojoki S, Bellomo C, Kareinen L, Andersson LC, et al. Hantavirus infection-induced B cell activation elevates free light chains levels in circulation. PLoS Pathog 2021; 17. doi:10.1371/journal.ppat.1009843

55. Koma T, Yoshimatsu K, Nagata N, Sato Y, Shimizu K, Yasuda SP, et al. Neutrophil Depletion Suppresses Pulmonary Vascular Hyperpermeability and Occurrence of Pulmonary Edema Caused by Hantavirus Infection in C.B-17 SCID Mice. J Virol 2014; 88: 7178–88. doi:10.1128/JVI.00254-14

56. Raftery MJ, Lalwani P, Krautkrӓmer E, Peters T, Scharffetter-Kochanek K, Krüger R, et al. β2 integrin mediates hantavirus-induced release of neutrophil extracellular traps. J Exp Med 2014; 211: 1485–97. doi:10.1084/jem.20131092

57. Strandin T, Mäkelä S, Mustonen J, Vaheri A. Neutrophil Activation in Acute Hemorrhagic Fever With Renal Syndrome Is Mediated by Hantavirus-Infected Microvascular Endothelial Cells. Front Immunol 2018; 9: 2098. doi:10.3389/fimmu.2018.02098

58. Marsac D, García S, Fournet A, Aguirre A, Pino K, Ferres M, et al. Infection of human monocyte-derived dendritic cells by ANDES Hantavirus enhances pro-inflammatory state, the secretion of active MMP-9 and indirectly enhances endothelial permeability. Virol J 2011; 8. doi:10.1186/1743-422X-8-223

59. Scholz S, Baharom F, Rankin G, Maleki KT, Gupta S, Vangeti S, et al. Human hantavirus infection elicits pronounced redistribution of mononuclear phagocytes in peripheral blood and airways. PLoS Pathog 2017; 13. doi:10.1371/journal.ppat.1006462

60. Raftery MJ, Lalwani P, Lütteke N, Kobak L, Giese T, Ulrich RG, et al. Replication in the Mononuclear Phagocyte System (MPS) as a Determinant of Hantavirus Pathogenicity. Front Cell Infect Microbiol 2020; 10: 281. doi:10.3389/fcimb.2020.00281

61. Vangeti S, Strandin T, Liu S, Tauriainen J, Raïsänen-Sokolowski A, Cabreraid L, et al. Monocyte subset redistribution from blood to kidneys in patients with Puumala virus caused hemorrhagic fever with renal syndrome. PLoS Pathog 2021; 17. doi:10.1371/journal.ppat.1009400

62. Cabrera LE, Schmotz C, Saleem MA, Lehtonen S, Vapalahti O, Vaheri A, et al. Increased Heparanase Levels in Urine during Acute Puumala Orthohantavirus Infection Are Associated with Disease Severity. Viruses 2022; 14: 450. doi:10.3390/v14030450

63. Niskanen S, Jääskeläinen A, Vapalahti O, Sironen T. Evaluation of real-time RT-PCR for diagnostic use in detection of puumala virus. Viruses 2019; 11. doi:10.3390/v11070661

64. Kyriakidis I, Papa A. Serum TNF-α, sTNFR1, IL-6, IL-8 and IL-10 levels in hemorrhagic fever with renal syndrome. Virus Res 2013; 175: 91–4. doi:10.1016/j.virusres.2013.03.020

65. Khaiboullina SF, Levis S, Morzunov SP, Martynova E V., Anokhin VA, Gusev OA, et al. Serum cytokine profiles differentiating hemorrhagic fever with renal syndrome and hantavirus pulmonary syndrome. Front Immunol 2017; 8. doi:10.3389/fimmu.2017.00567

66. Angulo J, Martínez-Valdebenito C, Marco C, Galeno H, Villagra E, Vera L, et al. Serum levels of interleukin-6 are linked to the severity of the disease caused by Andes Virus. PLoS Negl Trop Dis 2017; 11: e0005757. doi:10.1371/journal.pntd.0005757

67. Guo J, Guo X, Wang Y, Tian F, Luo W, Zou Y. Cytokine response to Hantaan virus infection in patients with hemorrhagic fever with renal syndrome. J Med Virol 2017; 89: 1139–45. doi:10.1002/jmv.24752

68. Walker JA, McKenzie ANJ. TH2 cell development and function. Nat Rev Immunol 2018; 18: 121–33. doi:10.1038/nri.2017.118

69. Kato A. Group 2 Innate Lymphoid Cells in Airway Diseases. Chest 2019; 156: 141–9. doi:10.1016/j.chest.2019.04.101

70. Ngo VL, Abo H, Maxim E, Harusato A, Geem D, Medina-Contreras O, et al. A cytokine network involving IL-36γ, IL-23, and IL-22 promotes antimicrobial defense and recovery from intestinal barrier damage. Proc Natl Acad Sci U S A 2018; 115: E5076–85. doi:10.1073/pnas.1718902115

71. Yudanin NA, Schmitz F, Flamar AL, Thome JJC, Tait Wojno E, Moeller JB, et al. Spatial and Temporal Mapping of Human Innate Lymphoid Cells Reveals Elements of Tissue Specificity. Immunity 2019; 50: 505–519.e4. doi:10.1016/j.immuni.2019.01.012

72. Gupta S, Braun M, Tischler ND, Stoltz M, Sundström KB, Björkström NK, et al. Hantavirus-infection Confers Resistance to Cytotoxic Lymphocyte-Mediated Apoptosis. Heise MT, editor. PLoS Pathog 2013; 9: e1003272. doi:10.1371/journal.ppat.1003272

73. Braun M, Björkström NK, Gupta S, Sundström K, Ahlm C, Klingström J, et al. NK Cell Activation in Human Hantavirus Infection Explained by Virus-Induced IL-15/IL15Rα Expression. Basler CF, editor. PLoS Pathog 2014; 10: e1004521. doi:10.1371/journal.ppat.1004521

74. Dhariwal J, Cameron A, Wong E, Paulsen M, Trujillo-Torralbo MB, Del Rosario A, et al. Pulmonary innate lymphoid cell responses during rhinovirus-induced asthma exacerbations in vivo: A clinical trial. Am J Respir Crit Care Med 2021; 204: 1259–73. doi:10.1164/rccm.202010-3754OC

75. Khaiboullina SF, Martynova E V, Khamidullina ZL, Lapteva E V, Nikolaeva I V, Anokhin V V, et al. Upregulation of IFN-γ and IL-12 is associated with a milder form of hantavirus hemorrhagic fever with renal syndrome. Eur J Clin Microbiol Infect Dis 2014; 33: 2149–56. doi:10.1007/s10096-014-2176-x

76. Peters CJ, Simpson GL, Levy H. Spectrum of hantavirus infection: hemorrhagic fever with renal syndrome and hantavirus pulmonary syndrome. Annu Rev Med 1999; 50: 531–45. doi:10.1146/annurev.med.50.1.531

77. Mustonen J, Mäkelä S, Outinen T, Laine O, Jylhävä J, Arstila PT, et al. The pathogenesis of nephropathia epidemica: New knowledge and unanswered questions. Antiviral Res 2013; 100: 589–604. doi:10.1016/j.antiviral.2013.10.001

78. Jiang H, Zheng X, Wang L, Du H, Wang P, Bai X. Hantavirus infection: a global zoonotic challenge. Virol Sin 2017; 32: 32–43. doi:10.1007/s12250-016-3899-x

79. Korva M, Saksida A, Kejžar N, Schmaljohn C, Avšič-Županc T. Viral load and immune response dynamics in patients with haemorrhagic fever with renal syndrome. Clin Microbiol Infect 2013; 19: E358–66. doi:10.1111/1469-0691.12218

80. Pettersson L, Thunberg T, Rocklöv J, Klingström J, Evander M, Ahlm C. Viral load and humoral immune response in association with disease severity in Puumala hantavirus-infected patients-implications for treatment. Clin Microbiol Infect 2014; 20: 235–41. doi:10.1111/1469-0691.12259

81. Mjösberg JM, Trifari S, Crellin NK, Peters CP, Van Drunen CM, Piet B, et al. Human IL-25-and IL-33-responsive type 2 innate lymphoid cells are defined by expression of CRTH2 and CD161. Nat Immunol 2011; 12: 1055–62. doi:10.1038/ni.2104

82. Mindt BC, Fritz JH, Duerr CU. Group 2 innate lymphoid cells in pulmonary immunity and tissue homeostasis. Front Immunol 2018; 9: 1. doi:10.3389/fimmu.2018.00840

83. Fonseca W, Lukacs NW, Elesela S, Malinczak CA. Role of ILC2 in Viral-Induced Lung Pathogenesis. Front Immunol 2021; 12. doi:10.3389/fimmu.2021.675169

84. Rodriguez-Rodriguez N, Gogoi M, McKenzie ANJ. Group 2 Innate Lymphoid Cells: Team Players in Regulating Asthma. Annu Rev Immunol 2021; 39: 167–98. doi:10.1146/annurev-immunol-110119-091711

85. Zhang Y, Zhang C, Zhuang R, Ma Y, Zhang Y, Yi J, et al. IL-33/ST2 Correlates with Severity of Haemorrhagic Fever with Renal Syndrome and Regulates the Inflammatory Response in Hantaan Virus-Infected Endothelial Cells. PLoS Negl Trop Dis 2015; 9. doi:10.1371/journal.pntd.0003514

86. Habtezion A, Nguyen LP, Hadeiba H, Butcher EC. Leukocyte Trafficking to the Small Intestine and Colon. Gastroenterology 2016; 150: 340–54. doi:10.1053/j.gastro.2015.10.046

87. Lee M, Kiefel H, Lajevic MD, Macauley MS, O’Hara E, Pan J, et al. Transcriptional programs of lymphoid tissue capillary and high endothelium reveal control mechanisms for lymphocyte homing. Nat Immunol 2014; 15: 982. doi:10.1038/NI.2983

88. Ito T, Carson WF, Cavassani KA, Connett JM, Kunkel SL. CCR6 as a mediator of immunity in the lung and gut. Exp Cell Res 2011; 317: 613–9. doi:10.1016/j.yexcr.2010.12.018

89. Bargatze RF, Jutila MA, Butcher EC. Distinct roles of L-selectin and integrins α4β7 and LFA-1 in lymphocyte homing to Peyer’s patch-HEV in situ: The multistep model confirmed and refined. Immunity 1995; 3: 99–108. doi:10.1016/1074-7613(95)90162-0

90. Solà-Riera C, Gupta S, Maleki KT, González-Rodriguez P, Saidi D, Zimmer CL, et al. Hantavirus Inhibits TRAIL-Mediated Killing of Infected Cells by Downregulating Death Receptor 5. Cell Rep 2019; 28: 2124–2139.e6. doi:10.1016/j.celrep.2019.07.066

91. Solà-Riera C, García M, Ljunggren HG, Klingström J. Hantavirus inhibits apoptosis by preventing mitochondrial membrane potential loss through up-regulation of the pro-survival factor BCL-2. PLoS Pathog 2020; 16: e1008297. doi:10.1371/journal.ppat.1008297

92. Katze MG, He Y, Gale M. Viruses and interferon: A fight for supremacy. Nat Rev Immunol 2002; 2: 675–87. doi:10.1038/nri888

93. Samuel CE. Antiviral actions of interferons. Clin Microbiol Rev 2001; 14: 778–809. doi:10.1128/CMR.14.4.778-809.2001

94. Kang S, Brown HM, Hwang S. Direct antiviral mechanisms of interferon-gamma. Immune Netw 2018; 18. doi:10.4110/in.2018.18.e33

95. Kak G, Raza M, Tiwari BK. Interferon-gamma (IFN-γ): Exploring its implications in infectious diseases. Biomol Concepts 2018; 9: 64–79. doi:10.1515/bmc-2018-0007

96. Kurioka A, Cosgrove C, Simoni Y, van Wilgenburg B, Geremia A, Björkander S, et al. CD161 defines a functionally distinct subset of pro-inflammatory natural killer cells. Front Immunol 2018; 9: 486. doi:10.3389/fimmu.2018.00486

97. Alter G, Jost S, Rihn S, Reyor LL, Nolan BE, Ghebremichael M, et al. Reduced frequencies of NKp30+NKp46+, CD161+, and NKG2D+ NK cells in acute HCV infection may predict viral clearance. J Hepatol 2011; 55: 278–88. doi:10.1016/j.jhep.2010.11.030

98. Petitdemange C, Becquart P, Wauquier N, Béziat V, Debré P, Leroy EM, et al. Unconventional repertoire profile is imprinted during acute chikungunya infection for natural killer cells polarization toward cytotoxicity. PLoS Pathog 2011; 7. doi:10.1371/journal.ppat.1002268

99. Leeansyah E, Ganesh A, Quigley MF, Sönnerborg A, Andersson J, Hunt PW, et al. Activation, exhaustion, and persistent decline of the antimicrobial MR1-restricted MAIT-cell population in chronic HIV-1 infection. Blood 2013; 121: 1124–35. doi:10.1182/blood-2012-07-445429

100. Wang Z, Guan D, Huo J, Biswas SK, Huang Y, Yang Y, et al. IL-10 Enhances Human Natural Killer Cell Effector Functions via Metabolic Reprogramming Regulated by mTORC1 Signaling. Front Immunol 2021; 12: 366. doi:10.3389/fimmu.2021.619195

