## Supplementary Material for "Innate lymphoid cells are activated and their levels correlate with viral load in patients with Puumala hantavirus caused hemorrhagic fever with renal syndrome"

**SUPPORTING INFORMATION**

**Table S1. Extended clinical and laboratory characteristics of HFRS patients.**

**Table S2. Antibodies and reagents used in flow cytometry.**

**S1. Correlations of clinical parameters and soluble factors in plasma of HFRS patients.**

**(a)** Level of soluble factors in plasma of HFRS patients in acute (n=15), early convalescence (n=16), and late convalescence (n=17) phase measured by multiplex immunoassay. Abbreviations: IL: interleukin. Graphs show data of individual subjects (circles) and the median (bars) ± interquartile range. Statistical significance was assessed using the Wilcoxon signed-rank test. Severe patients are indicated by a black circle. **(b)** Spearman correlation matrix of the clinical parameters and the soluble markers measured in plasma of acute HFRS patients by a multiplex immunoassay. The colour of the circles indicates positive (red) and negative (blue) correlations that were statistically significant ($p < 0.05$) as measured by the Spearman's rank correlation coefficient test. The colour intensity and the size of the circle are proportional to the correlation coefficients.

**S2. Gating strategy for ILCs and NK identification in flow cytometry and UMAP analysis.**

Gating strategy used for the identification of total ILCs and NK cells by flow cytometry.

**S3. NK cell subsets are activated, proliferative and present a migratory profile in peripheral blood of HFRS patients**.

**(a-b)** Percentage of CD69$^{+}$, Ki-67$^{+}$, HLA-DR$^{+}$, NKG2A$^{+}$, NKp44$^{+}$, CCR6$^{+}$, CCR10$^{+}$, α4β7$^{+}$, CD45RA$^{+}$, and CD161$^{+}$ **(a)** CD56$^{dim}$ NK cells and **(b)** CD56$^{bright}$ NK cells in control donors (n=10) and HFRS patients during the acute (n=15), early convalescence (n=16), and late convalescence (n=17) phase.

Bar graphs are shown as mean and lines connect paired samples from the same patient. Statistical significance was assessed using the Wilcoxon signed-rank test to compare groups of HFRS patients, and the Kruskal-Wallis test followed by Dunn's multiple comparisons test to compare healthy controls with groups of HFRS patients. Severe patients are indicated by a black circle. *$p < 0.05$; **$p < 0.01$; ***$p < 0.001$; ****$p < 0.0001$.

**S4. Correlations of soluble factors and clinical parameters with NK cells in HFRS patients.**

Spearman correlation matrix of the level of soluble factors in plasma and the percentage of **(a)** NK cells, **(b)** $CD56^{dim}$ NK cells, and **(c)** $CD56^{bright}$ NK cells in acute HFRS patients. Spearman correlation matrix of the clinical parameters and the percentage of **(d)** NK cells, **(e)** $CD56^{dim}$ NK cells, and **(f)** $CD56^{bright}$ NK cells in acute HFRS patients. The colour of the circles indicates positive (red) and negative (blue) correlations that were statistically significant ($p < 0.05$) as measured by the Spearman's rank correlation coefficient test. The colour intensity and the size of the circle are proportional to the correlation coefficients. Ly: lymphocytes. Days a. symp.: days after symptoms onset.

**S5. ILCs display an activated and proliferative profile in peripheral blood of HFRS patients.**

**(a) i.** Percentage of $CD69^+$, Ki-$67^+$, HLA-$DR^+$, $NKp44^+$, $CD56^+$, $CCR6^+$, $CCR10^+$, $\alpha4\beta7^+$ and $CD45RA^+$ ILCs in control donors (n=10) and HFRS patients during the acute (n=15), early convalescence (n=16), and late convalescence (n=17) phase.

**(b)** Spearman rank correlation between plasma IL-10 levels and the percentage of $CD69^+$ ILC, plasma CCL27 levels and the percentage of $CCR10^+$ ILC, and plasma IL-7 levels and the percentage of Ki-$67^+$ ILC in acute HFRS patients (n = 15).

Bar graphs are shown as mean and lines connect paired samples from the same patient. Statistical significance was assessed using the Wilcoxon signed-rank test to compare groups of HFRS patients, and the Kruskal-Wallis test followed by Dunn's multiple comparisons test to compare healthy controls with groups of HFRS patients. Severe patients are indicated by a black circle.

Severe patients are indicated by a black circle. *p < 0.05; **p < 0.01; ***p < 0.001; ****p < 0.0001.

**S6. Correlations of soluble factors and clinical parameters with ILCs in HFRS patients.**

Spearman correlation matrix of the clinical parameters and the percentage of **(a)** ILCs, **(b)** ILC2, and **(c)** nILC in acute HFRS patients. Spearman correlation matrix of the clinical parameters and the percentage of **(d)** ILCs, **(e)** ILC2, and **(f)** nILC in acute HFRS patients. The colour of the circles indicates positive (red) and negative (blue) correlations that were statistically significant (p < 0.05) as measured by the Spearman's rank correlation coefficient test. The colour intensity and the size of the circle are proportional to the correlation coefficients. Ly: lymphocytes. Days a. symp.: days after symptoms onset.

**Supplementary table I.** Antibodies and reagents used in flow cytometry

| Laser | Filter | Fluorochrome | Marker | Clone | Company | Catalog number | RRID |
|---|---|---|---|---|---|---|---|
| UV | | BUV395 | CD45 | HI30 | BD | 563791 | AB_2744400 |
| | | BUV737 | CCR6 | 11A9 | BD | 612780 | AB_2870109 |
| 488 | 530/30 | FITC | CD1a | HI149 | Biolegend | 300104 | AB_314018 |
| | | | CD14 | Tuk4 | Life Technologies | MHCD14014 | AB_1464899 |
| | | | CD19 | 4G7 | BD Biosciences | 345776 | AB_2868804 |
| | | | CD34 | 581 | Biolegend | 343504 | AB_1731852 |
| | | | CD123 | 6H6 | Biolegend | 306014 | AB_2124259 |
| | | | BDCA2 | AC144 | Miltenyi | 130-113-192 | AB_2726017 |
| | | | FcER1 | AER-37 (CRA1) | Biolegend | 334608 | AB_1227653 |
| | | | TCRab | IP26 | Biolegend | 306706 | AB_314644 |
| | | | TCRgd | B1 | Biolegend | 331208 | AB_1575108 |
| 639 | 670/30 | APC | NKG2A | Z199 | Beckman Coulter | A60797 | AB_10643105 |
| | 730/45 | A700 | Ki-67 | B56 | BD | 561277 | AB_10611571 |
| 405 | 450/50 | BV 421 | CCR10 | 1B5 | BD | 564770 | AB_2738942 |
| | 525/50 | BV510 | CD69 | FN50 | Biolegend | 747521 | N/A |
| | 585/42 | BV 570 | CD3 | UCHT1 | Biolegend | 300436 | AB_2562124 |
| | 610/20 | BV605 | CD161 | HP-3810 | Biolegend | 339916 | AB_2563607 |
| | 670/30 | BV 650 | α4β7 biotin | HU117 | R&D | MAB10078-100 | N/A |
| | | | streptavidin | | Biolegend | 405231 | N/A |
| | 710/50 | BV 711 | CD56 | HCD56 | Biolegend | 318336 | AB_2562417 |
| | 780/60 | BV 785 | CD45RA | HI100 | Biolegend | 304140 | AB_2563816 |
| 561 | 586/15 | PE | HLADR | L243 | Biolegend | 307606 | AB_314684 |
| | 620/14 | PE-Dazzle 594 | CRTH2 | BM16 | Biolegend | 350125 | AB_2572052 |
| | 661/20 | PE-Cy5 | NKp44 | Z231 | Beckman Coulter | A66903 | N/A |
| | 710/50 | PE-Cy5.5 | CD117 | 104D2D1 | Beckman Coulter | B96754 | N/A |
| | 780/60 | PE-Cy7 | CD127 | R34.34 | Beckman Coulter | A64618 | AB_2833031 |

**Supplementary table 2**. Extended clinical and laboratory characteristics of HFRS patients

| Patient | Age range (years) | Gender | Days after symptoms onset | Days after hospitalization | Days hospitalized | Viral load (PUUV S RNA copies/mL) | Leucocyte count (10^9/L) | Hematocrit (L/L) | Platelet count (10^9/L) | CRP (mg/L) | Creatinine (µmol/L) | CMV (copies/mL) | EBV (copies/mL) | MAP | Severity score | Severity |
|---|---|---|---|---|---|---|---|---|---|---|---|---|---|---|---|---|
| P01 | 56-60 | F | 7 | 1 | 6 | 187975 | 9 | 0.35 | 114 | 81.7 | 54 | neg | neg | 82 | 1 | Mild |
| P03 | 46-50 | M | 6 | 2 | 9 | 88955 | 21.8 | 0.44 | 91 | 87.3 | 519 | neg | 338 | 92 | 6 | Severe |
| P04 | 26-30 | F | 9 | 4 | 5 | 71901 | 8.8 | 0.36 | 131 | 43.1 | 191 | neg | neg | 74 | 3 | Mild |
| P05 | 36-40 | M | 6 | 1 | 2 | 1867667 | 10.3 | 0.44 | 52 | 111.2 | 120 | neg | neg | 92 | 3 | Mild |
| P06 | 51-55 | M | 6 | 1 | 4 | 94272 | 12.2 | 0.46 | 89 | 27 | 179 | neg | neg | 83 | 4 | Mild |
| P07 | 41-45 | M | 6 | 1 | 4 | 149025 | 8.2 | 0.36 | 121 | 94 | 498 | neg | neg | 82 | 5 | Severe |
| P08 | 26-30 | F | 8 | 3 | 5 | 70523 | 7.5 | 0.38 | 102 | 20 | 85 | neg | neg | 72 | 1 | Mild |
| P09 | 21-25 | F | 8 | 3 | NA | 112205 | 18.2 | 0.36 | 86 | 58.2 | 107 | neg | neg | 68 | 3 | Mild |
| P10 | 66-70 | F | 6 | 1 | 9 | 1143969 | 10.8 | 0.34 | 118 | 33.5 | 431 | neg | neg | 76 | 4 | Mild |
| P11 | 51-55 | F | 6 | 2 | 7 | ND | 12 | 0.37 | 82 | 33.4 | 254 | neg | neg | 98 | 4 | Mild |
| P13 | 26-30 | F | 7 | 3 | 6 | 47300 | 6.9 | 0.35 | 66 | 94 | 155 | neg | neg | 91 | 3 | Mild |
| P14 | 26-30 | F | 8 | 2 | 4 | ND | 5.7 | 0.34 | 199 | 24.7 | 137 | neg | neg | 100 | 1 | Mild |
| P15 | 31-35 | F | 5 | 2 | 5 | 181000 | NA | NA | NA | 93.5 | 42 | neg | neg | 83 | 0 | Mild |
| P16 | 26-30 | F | 6 | 2 | 4 | 261000 | 8.6 | 0.4 | 115 | 51.8 | NA | neg | neg | 76 | 1 | Mild |
| P17 | 31-35 | M | 7 | 3 | NA | 41600 | 9.5 | 0.37 | 86 | 78.5 | 99 | neg | neg | 90 | 2 | Mild |

Data in the table corresponds to the HFRS patients in acute phase.
F = female, M = male, NA: not available; ND: not detectable; neg: negative; CMV: Cytomegalovirus; EBV: Epstein-Barr virus.
Platelet count; normal range 150–360 x10^9/L.
Plasma C-reactive protein (CRP); reference <3 mg/L.
Plasma creatinine; reference <90 µmol/L for women, <100 µmol/L for men.
Mean arterial blood pressure (MAP); reference 70–100 mmHg.

**Figure S1**

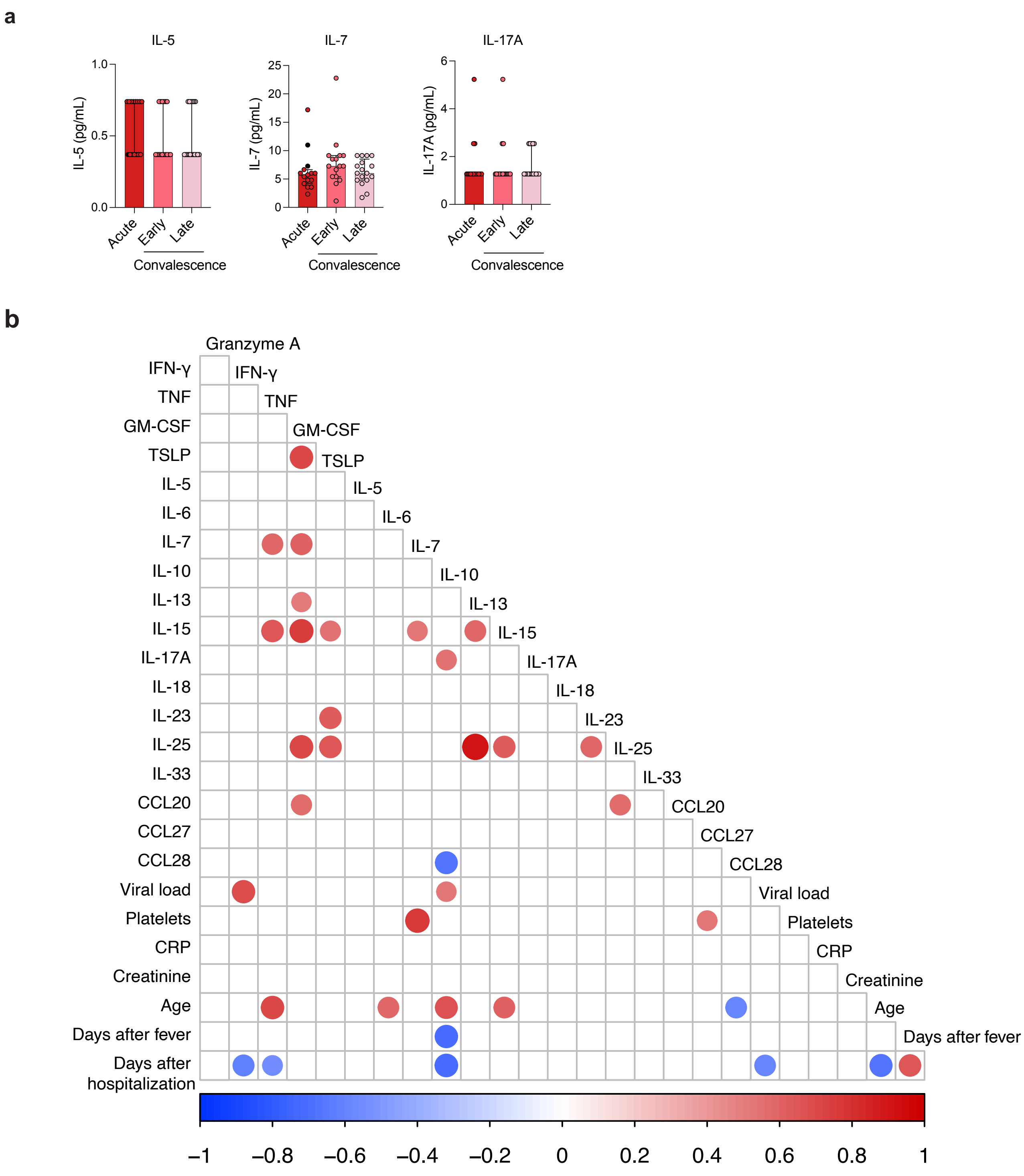
a
IL-5
IL-5 (pg/mL)
1.0
0.5
0.0
Acute
Early
Late
Convalescence
IL-7
IL-7 (pg/mL)
25
20
15
10
5
0
IL-17A
IL-17A (pg/mL)
6
4
2
0
b
Granzyme A
IFN-γ
TNF
GM-CSF
TSLP
IL-5
IL-6
IL-7
IL-10
IL-13
IL-15
IL-17A
IL-18
IL-23
IL-25
IL-33
CCL20
CCL27
CCL28
Viral load
Platelets
CRP
Creatinine
Age
Days after fever
Days after hospitalization
–1
–0.8
–0.6
–0.4
–0.2
0
0.2
0.4
0.6
0.8
1

**Figure S2**

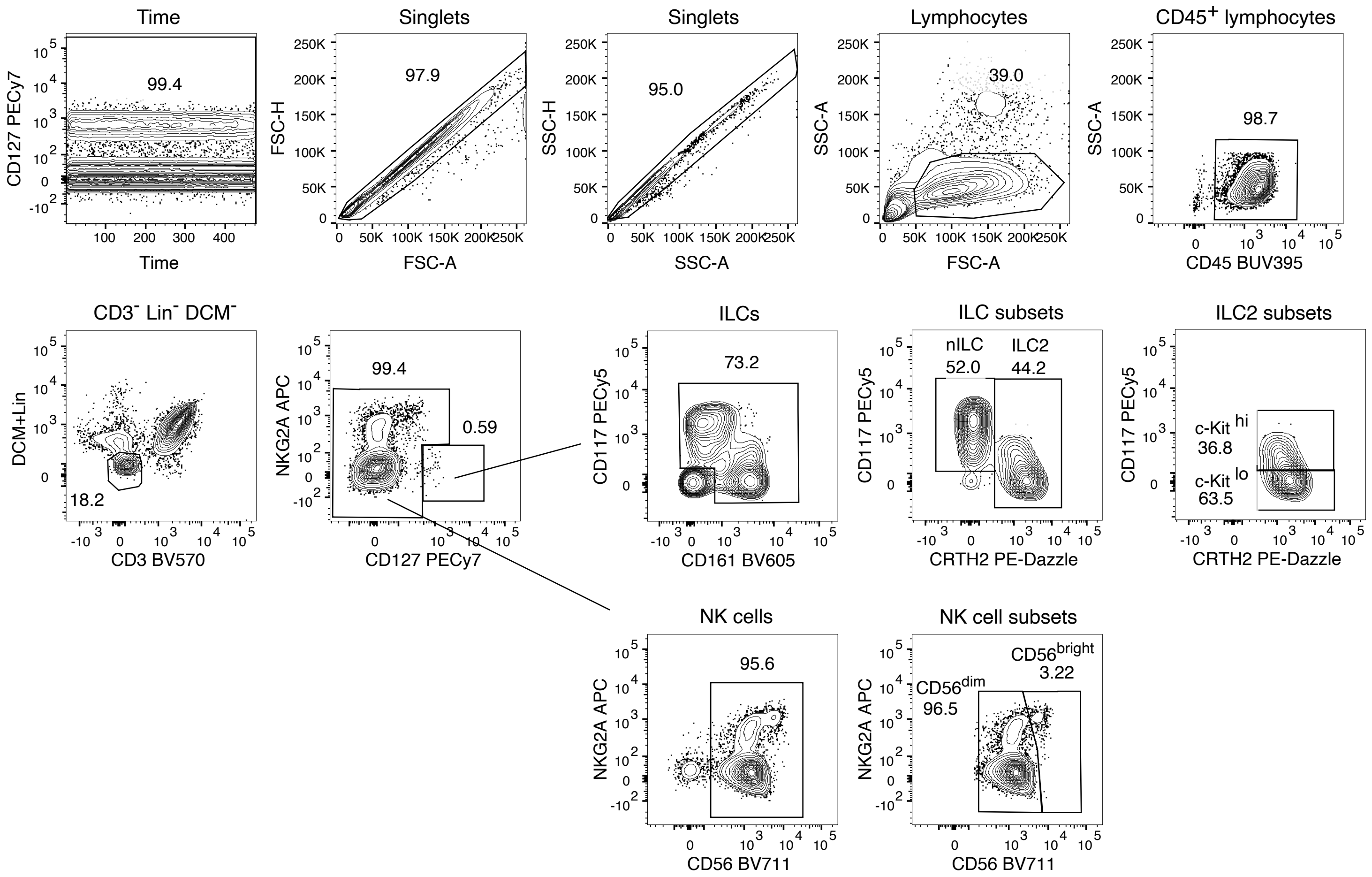

Time
99.4
CD127 PECy7
Time
Singlets
97.9
FSC-H
FSC-A
Singlets
95.0
SSC-H
SSC-A
Lymphocytes
39.0
SSC-A
FSC-A
CD45+ lymphocytes
98.7
SSC-A
CD45 BUV395
CD3- Lin- DCM-
18.2
DCM+Lin
CD3 BV570
99.4
0.59
NKG2A APC
CD127 PECy7
ILCs
73.2
CD117 PECy5
CD161 BV605
ILC subsets
nILC
52.0
ILC2
44.2
CD117 PECy5
CRTH2 PE-Dazzle
ILC2 subsets
c-Kit hi
36.8
c-Kit lo
63.5
CD117 PECy5
CRTH2 PE-Dazzle
NK cells
95.6
NKG2A APC
CD56 BV711
NK cell subsets
CD56bright
3.22
CD56dim
96.5
NKG2A APC
CD56 BV711

### Figure S3

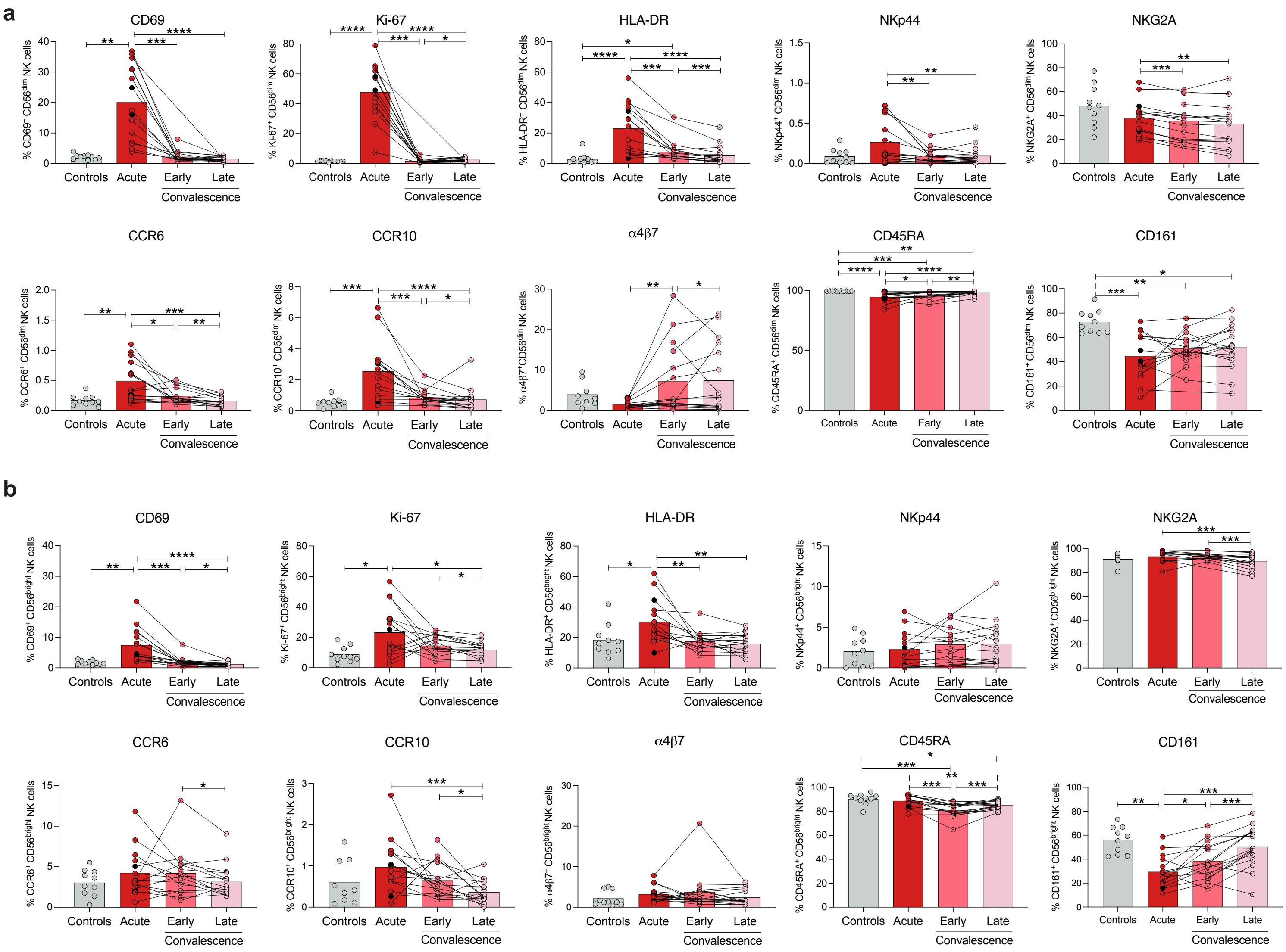
a
CD69
Ki-67
HLA-DR
NKp44
NKG2A
CCR6
CCR10
α4β7
CD45RA
CD161
% CD69+ CD56dim NK cells
% Ki-67+ CD56dim NK cells
% HLA-DR+ CD56dim NK cells
% NKp44+ CD56dim NK cells
% NKG2A+ CD56dim NK cells
% CCR6+ CD56dim NK cells
% CCR10+ CD56dim NK cells
% α4β7+ CD56dim NK cells
% CD45RA+ CD56dim NK cells
% CD161+ CD56dim NK cells
Controls
Acute
Early
Late
Convalescence
b
CD69
Ki-67
HLA-DR
NKp44
NKG2A
CCR6
CCR10
α4β7
CD45RA
CD161
% CD69+ CD56bright NK cells
% Ki-67+ CD56bright NK cells
% HLA-DR+ CD56bright NK cells
% NKp44+ CD56bright NK cells
% NKG2A+ CD56bright NK cells
% CCR6+ CD56bright NK cells
% CCR10+ CD56bright NK cells
% α4β7+ CD56bright NK cells
% CD45RA+ CD56bright NK cells
% CD161+ CD56bright NK cells
Controls
Acute
Early
Late
Convalescence

#### Figure S4

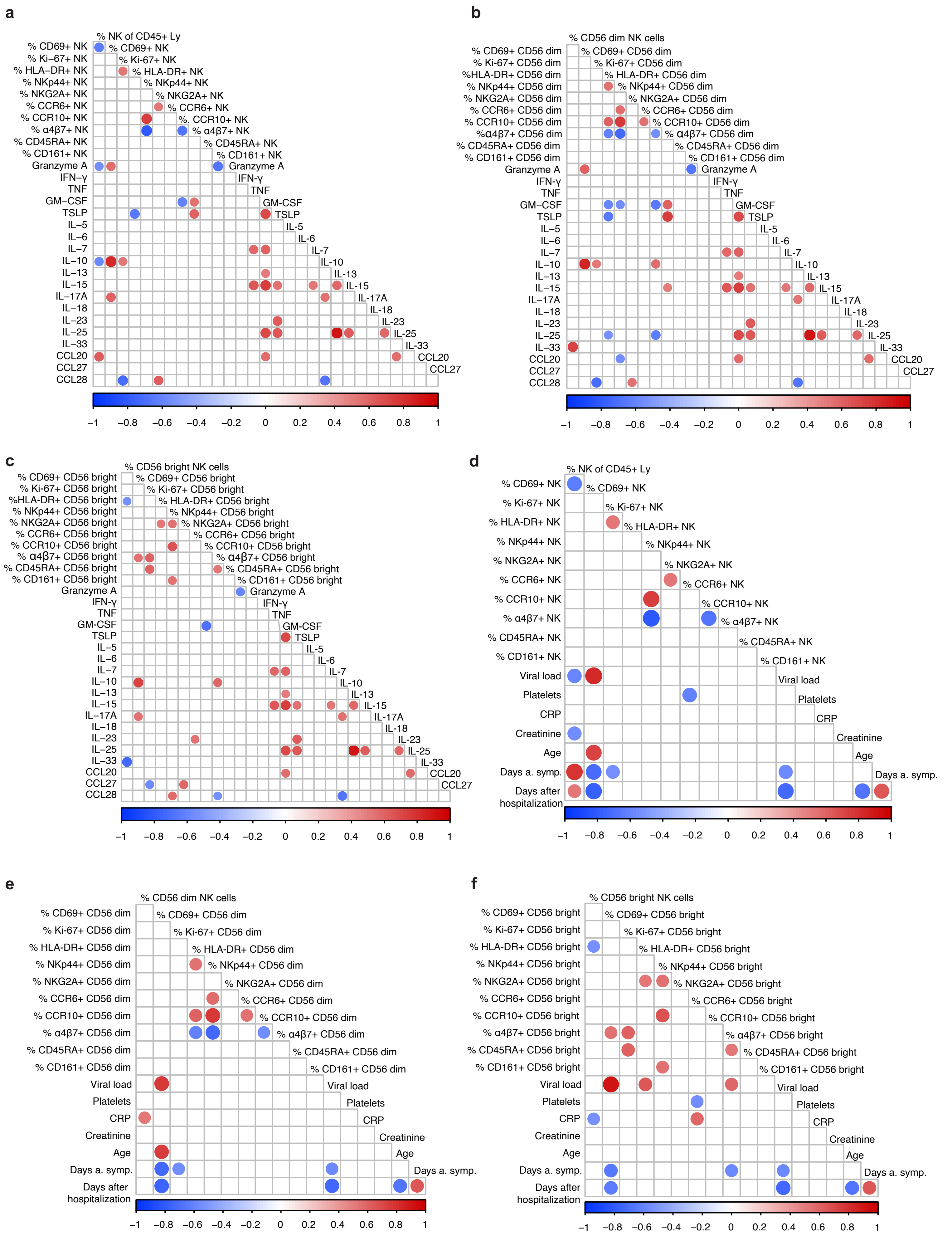
a
% NK of CD45+ Ly
% CD69+ NK
% Ki–67+ NK
% HLA–DR+ NK
% NKp44+ NK
% NKG2A+ NK
% CCR6+ NK
% CCR10+ NK
% α4β7+ NK
% CD45RA+ NK
% CD161+ NK
Granzyme A
IFN–γ
TNF
GM–CSF
TSLP
IL–5
IL–6
IL–7
IL–10
IL–13
IL–15
IL–17A
IL–18
IL–23
IL–25
IL–33
CCL20
CCL27
CCL28
–1 –0.8 –0.6 –0.4 –0.2 0 0.2 0.4 0.6 0.8 1
b
% CD56 dim NK cells
% CD69+ CD56 dim
% Ki-67+ CD56 dim
%HLA-DR+ CD56 dim
% NKp44+ CD56 dim
% NKG2A+ CD56 dim
% CCR6+ CD56 dim
% CCR10+ CD56 dim
%α4β7+ CD56 dim
% CD45RA+ CD56 dim
% CD161+ CD56 dim
c
% CD56 bright NK cells
% CD69+ CD56 bright
% Ki-67+ CD56 bright
%HLA-DR+ CD56 bright
% NKp44+ CD56 bright
% NKG2A+ CD56 bright
% CCR6+ CD56 bright
% CCR10+ CD56 bright
% α4β7+ CD56 bright
% CD45RA+ CD56 bright
% CD161+ CD56 bright
d
Viral load
Platelets
CRP
Creatinine
Age
Days a. symp.
Days after hospitalization
e
f

**Figure S5**

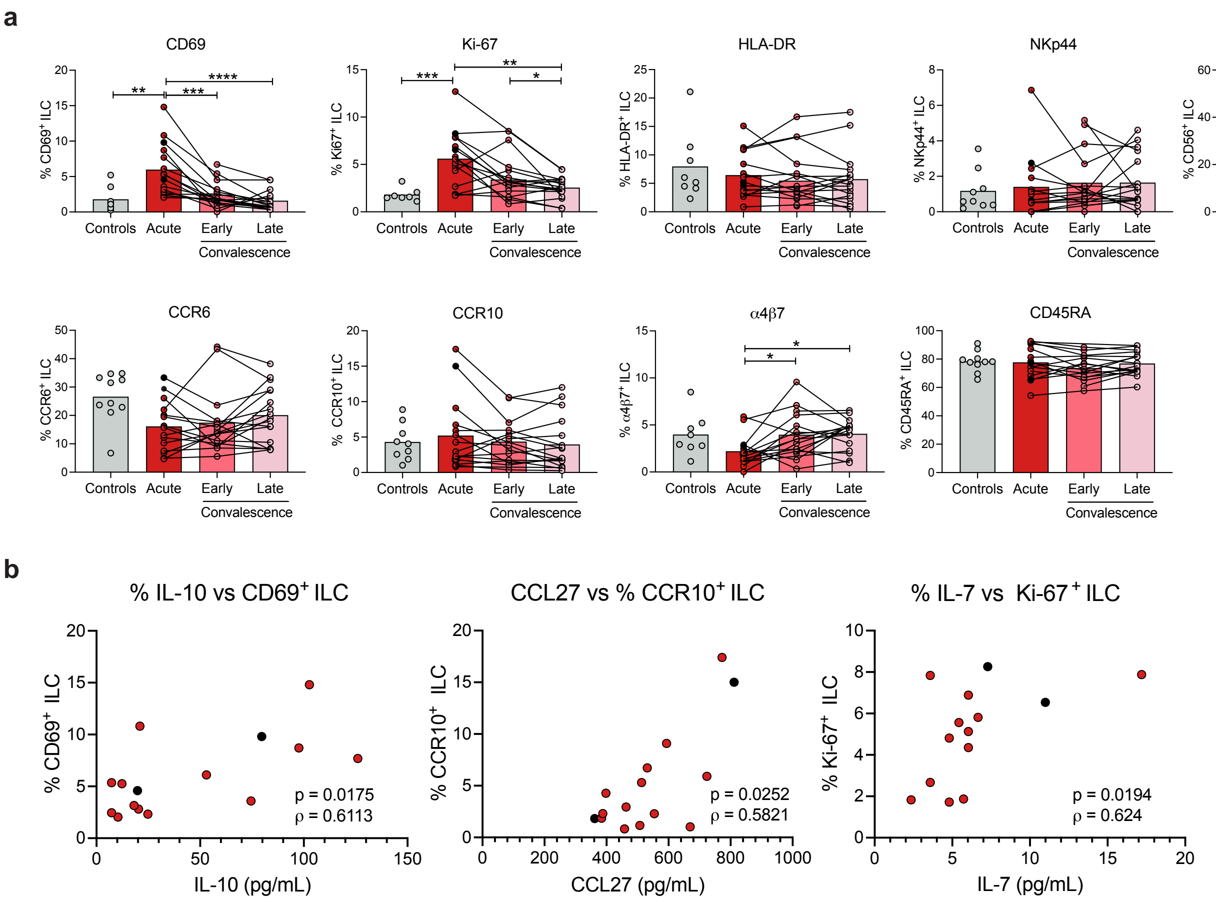
a
CD69
% CD69+ ILC
Ki-67
% Ki67+ ILC
HLA-DR
% HLA-DR+ ILC
NKp44
% NKp44+ ILC
CD56
% CD56+ ILC
CCR6
% CCR6+ ILC
CCR10
% CCR10+ ILC
α4β7
% α4β7+ ILC
CD45RA
% CD45RA+ ILC
Controls
Acute
Early
Late
Convalescence
b
% IL-10 vs CD69+ ILC
% CD69+ ILC
IL-10 (pg/mL)
p = 0.0175
ρ = 0.6113
CCL27 vs % CCR10+ ILC
% CCR10+ ILC
CCL27 (pg/mL)
p = 0.0252
ρ = 0.5821
% IL-7 vs Ki-67+ ILC
% Ki-67+ ILC
IL-7 (pg/mL)
p = 0.0194
ρ = 0.624

**Figure S6**

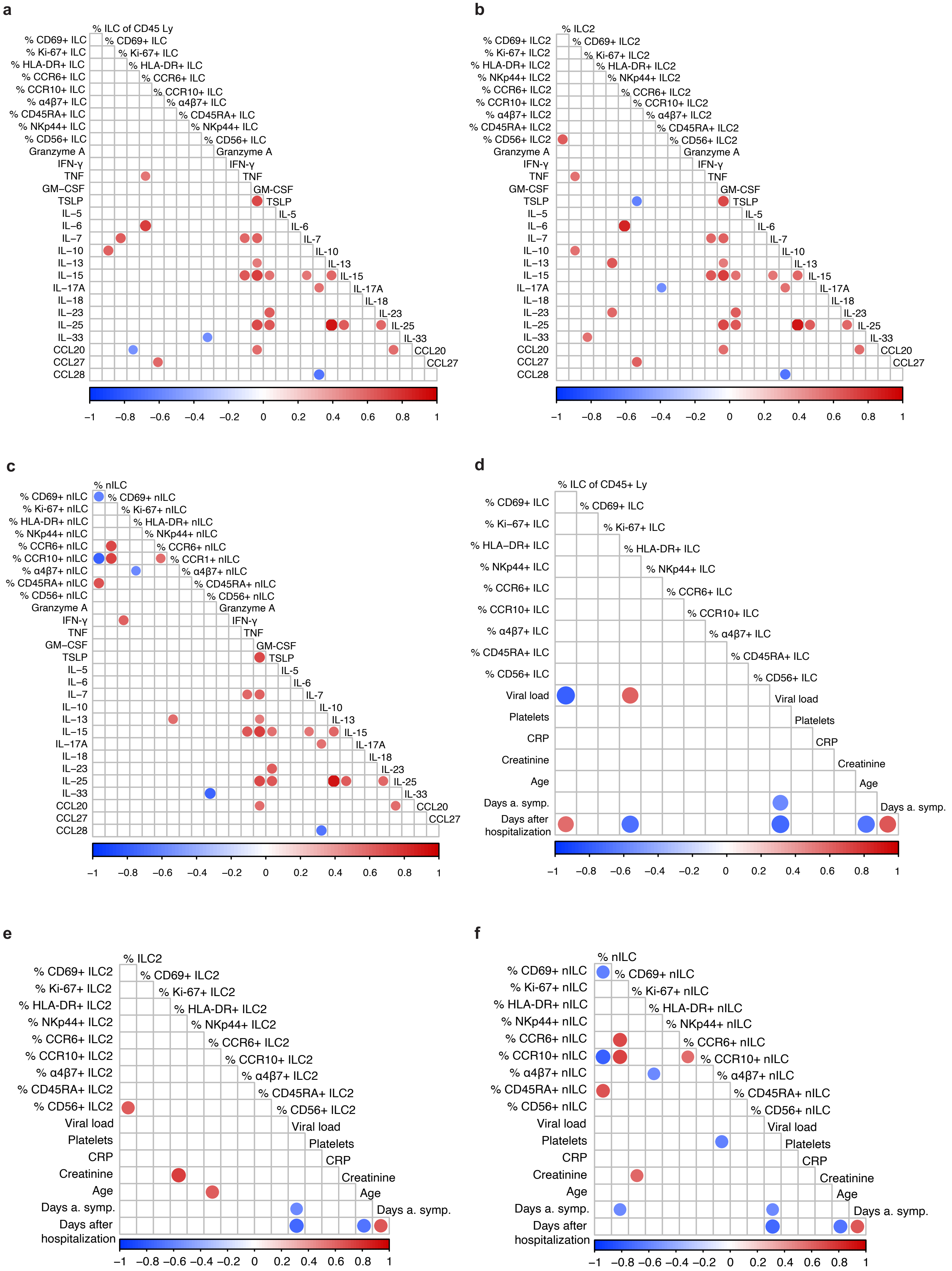

a
% ILC of CD45 Ly
% CD69+ ILC
% Ki-67+ ILC
% HLA-DR+ ILC
% CCR6+ ILC
% CCR10+ ILC
% α4β7+ ILC
% CD45RA+ ILC
% NKp44+ ILC
% CD56+ ILC
Granzyme A
IFN-γ
TNF
GM–CSF
TSLP
IL–5
IL–6
IL–7
IL–10
IL–13
IL–15
IL–17A
IL–18
IL–23
IL–25
IL–33
CCL20
CCL27
CCL28
–1 –0.8 –0.6 –0.4 –0.2 0 0.2 0.4 0.6 0.8 1
b
% ILC2
% CD69+ ILC2
% Ki-67+ ILC2
% HLA-DR+ ILC2
% NKp44+ ILC2
% CCR6+ ILC2
% CCR10+ ILC2
% α4β7+ ILC2
% CD45RA+ ILC2
% CD56+ ILC2
Granzyme A
IFN-γ
TNF
GM–CSF
TSLP
IL–5
IL–6
IL–7
IL–10
IL–13
IL–15
IL–17A
IL–18
IL–23
IL–25
IL–33
CCL20
CCL27
CCL28
–1 –0.8 –0.6 –0.4 –0.2 0 0.2 0.4 0.6 0.8 1
c
% nILC
% CD69+ nILC
% Ki-67+ nILC
% HLA-DR+ nILC
% NKp44+ nILC
% CCR6+ nILC
% CCR10+ nILC
% CCR1+ nILC
% α4β7+ nILC
% CD45RA+ nILC
% CD56+ nILC
Granzyme A
IFN-γ
TNF
GM–CSF
TSLP
IL–5
IL–6
IL–7
IL–10
IL–13
IL–15
IL–17A
IL–18
IL–23
IL–25
IL–33
CCL20
CCL27
CCL28
–1 –0.8 –0.6 –0.4 –0.2 0 0.2 0.4 0.6 0.8 1
d
% ILC of CD45+ Ly
% CD69+ ILC
% Ki–67+ ILC
% HLA–DR+ ILC
% NKp44+ ILC
% CCR6+ ILC
% CCR10+ ILC
% α4β7+ ILC
% CD45RA+ ILC
% CD56+ ILC
Viral load
Platelets
CRP
Creatinine
Age
Days a. symp.
Days after hospitalization
–1 –0.8 –0.6 –0.4 –0.2 0 0.2 0.4 0.6 0.8 1
e
% ILC2
% CD69+ ILC2
% Ki-67+ ILC2
% HLA-DR+ ILC2
% NKp44+ ILC2
% CCR6+ ILC2
% CCR10+ ILC2
% α4β7+ ILC2
% CD45RA+ ILC2
% CD56+ ILC2
Viral load
Platelets
CRP
Creatinine
Age
Days a. symp.
Days after hospitalization
–1 –0.8 –0.6 –0.4 –0.2 0 0.2 0.4 0.6 0.8 1
f
% nILC
% CD69+ nILC
% Ki-67+ nILC
% HLA-DR+ nILC
% NKp44+ nILC
% CCR6+ nILC
% CCR10+ nILC
% α4β7+ nILC
% CD45RA+ nILC
% CD56+ nILC
Viral load
Platelets
CRP
Creatinine
Age
Days a. symp.
Days after hospitalization
–1 –0.8 –0.6 –0.4 –0.2 0 0.2 0.4 0.6 0.8 1
